# Genomic surveillance of a deeply sampled local population reveals age-specific drivers of RSV transmission

**DOI:** 10.64898/2026.05.07.26350887

**Authors:** Jiye Kwon, Ellen M. de Vries, Philippe Lemey, Ke Li, Mallery Breban, Kelly Laing, Dave Ferguson, Wade Schulz, Carlos R. Oliveira, Louis J. Bont, Virginia E. Pitzer, Daniel M. Weinberger, Nathan D. Grubaugh, Verity Hill, Seth Redmond

## Abstract

Respiratory syncytial virus (RSV) disproportionately causes severe infections among infants and older adults, yet the key age group responsible for viral spread to other age groups remains poorly defined. While current immunization approaches effectively reduce disease severity among the most vulnerable, identifying the core drivers of infection is essential to effectively disrupt population-level transmission. By generating 910 whole-genome viral sequences of RSV from all age groups (<1 to 65+ years) in Connecticut, we identified that children aged 12-35 months are the primary drivers of viral transmission to other age groups. This group significantly shapes the genetic diversity of circulating strains. Furthermore, we found that RSV is introduced into the community through frequent and independent entries from other US regions throughout the year, rather than through a single explosive seasonal introduction or long-term local persistence. Ultimately, our findings justify prevention strategies that expand beyond reducing disease burden to actively prioritizing the reduction of transmission and infection.

## Introduction

Respiratory syncytial virus (RSV) remains the leading cause of lower respiratory tract infections globally, driving substantial morbidity and mortality(*1–4*). Although the established disease burden is highest among young children, immunocompromised individuals, and older adults, all age groups are susceptible to RSV reinfection throughout life (*5*). Since 2023, the U.S. Food and Drug Administration (FDA) has approved five new effective prophylactic products against RSV, including vaccines and long-acting monoclonal antibodies(*6–10*). These new options primarily aim to reduce disease severity among the most clinically vulnerable populations. Ultimately, transitioning from individual protection to optimal population-level control requires identifying the key drivers of transmission beyond protection of clinical disease.

Achieving sustained control requires resolving two critical, yet poorly understood, dimensions of RSV transmission. First, it remains unclear which specific age cohorts drive the RSV genetic diversity and transmission. Older siblings (broadly categorized as ∼1 to 15 years old) are frequently cited as the primary source of infant infections (*11*, *12*). However, current evidence relies heavily on within-household transmission studies(*11*, *12*), and there are scarce data from robust population-level studies with detailed individual-level age data to determine age-specific roles in transmission. Second, the mechanism of introduction and establishment in a local population remains unresolved. In the U.S., RSV epidemics typically begin in the southeast, peaking in late fall/early winter and then spreading through the Northeast and Midwest, as demonstrated by epidemiological studies(*13–16*). Furthermore, the timing of peaks in hospitalization varies by age group, with children under 5 years old experiencing peaks approximately one month earlier than older adults(*17*), the precise drivers behind this spatially and temporally structured pattern remain unresolved(*13*). Without a granular understanding of these regional and seasonal dynamics, despite the availability of new prophylactic tools, our ability to optimize prophylaxis administration or design clinical trials for population-level impact is stymied.

To answer these questions, genomic sequences and phylogenetic approaches provide a powerful framework for inferring the unobserved transmission dynamics that cannot be answered by epidemiological data alone(*18*). While phylogenetics allows us to reconstruct the evolutionary relationships between sequences, recent advances in phylodynamics have further expanded this toolkit (*18*). By integrating both genomic and non-genomic data—such as age and location information –phylodynamic approaches provide high-resolution insights into viral spread. Specifically, these analyses can disentangle the degree of introduction versus persistence(*19*), infer transmission directionality, and characterize inter-age-group dynamics(*20*). By leveraging local genomic surveillance to power this analytical framework, we can translate viral sequences into actionable public health insight. These insights provide the evidence to inform the design of targeted strategies and ensure the effective implementation of future vaccination programs. For instance, as the field establishes evidence around the effectiveness of current interventions in reducing RSV transmission or preventing infection, we can evaluate whether we should consider vaccinating other age groups that may contribute to broader transmission.

In this work, we present a high-resolution phylodynamic investigation of RSV lineage introduction and local transmission patterns across two seasons in Connecticut, U.S. Our dataset includes individual-level age information across two consecutive RSV seasons, providing comprehensive coverage of all age groups, from infants to older adults. The main objectives of our study are to: 1) identify the local Connecticut lineages relative to global diversity across both subtypes; 2) quantify and describe the degree of local lineage persistence through multiple seasons and the timing of introduction; and 3) compare virus transmission between age groups to identify the relative contributions of different age groups. Ultimately, this in-depth resolution of local transmission dynamics of RSV viral lineages offers a vital blueprint for optimizing and refining the design of future immunization programs across the United States.

## Results

### Local RSV dynamics reveal frequent, independent introductions of RSV into Connecticut

We established a hospital-based genomic surveillance system in Connecticut, utilizing an amplicon panel that targets both of the major RSV subtypes, A and B, and avoiding the need for prior subtyping(*21*). We generated a deeply sampled local population dataset of 910 whole-genome RSV sequences spanning two RSV seasons, from October 2, 2023, to February 19, 2025, with 415 classified as RSV-A and 495 classified as RSV-B (**Figure 1A**). Our data from Connecticut showed that the local 2023/24 season was dominated by RSV-B (81%), with a switch in 2024/25 to predominantly RSV-A (76%) (**Figure 1A**).

**Figure 1.**
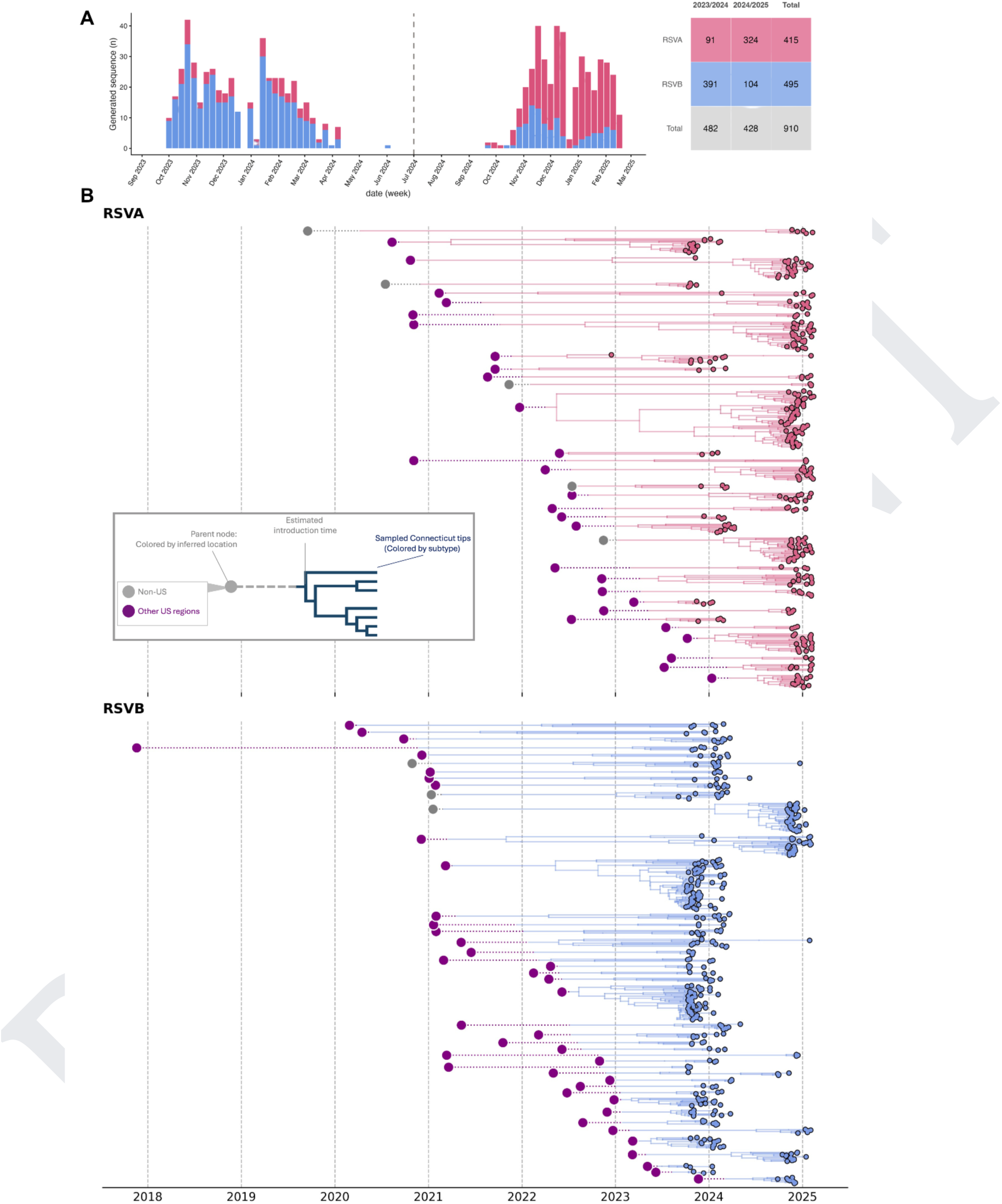
Connecticut respiratory syncytial virus (RSV) sequences. (A) Temporal distribution and contingency table of Connecticut RSV sequenced isolates colored by RSV season and subtype (n = 910). (B) Exploded trees colored by the inferred location of the parent of the most recent estimated introduction node inferred to be in Connecticut. Nodes are colored based on the inferred location. Grey nodes indicate an international parent node, while purple nodes indicate a U.S. location parent node. Branches are colored by RSV subtype.

We performed phylogeographic reconstruction of RSV-A and RSV-B independently to infer the timing of introduction and spatial connectivity of Connecticut lineages relative to sequences from other states and outside of the U.S. (**Supplemental Figure 1**). We found that distinct RSV-A and RSV-B clades circulating locally in Connecticut represent a broad sample of the global RSV genetic diversity. We identified 96 RSV-A and 138 RSV-B independent introduction events, of which 76 consisted of at least three sequences, indicating onward transmission within the state, which we refer to as “lineages”.

Our analysis demonstrated that these Connecticut lineages are more closely related to RSV sequences sampled in other U.S. states than they are to international sequences (**Figure 1B**). Only 8 (11%) of the Connecticut lineages showed a posterior probability of >0.5 supporting international location assignment for the origin of introduction, therefore we estimate that the remaining 89% were introduced from another U.S. region (**Figure 1B**).

We therefore established that RSV transmission relies on numerous separate introductions, mostly from other states in the U.S., thereby demonstrating that the local RSV activity is driven by high connectivity to the broader U.S. viral transmission.

### Local RSV lineage detection and persistence

We infer that RSV introductions (grey diamonds) occur year-round, across all subtypes (**Figure 2B**). Across both analyzed seasons, the optimal detection timing for new lineages was concentrated between October and January. The initial detection (blue triangle) of introduced local lineages preceded the peak of the epidemic season, with 81% of new lineages detected before January 1st, irrespective of their estimated phylogeographic introduction timing. However, the optimal timing for detecting newly introduced lineages is aligned with two concurrent events: the rise in RSV testing rates across all age groups (**Supplemental Figure 2**) and the characteristic seasonal increase in RSV hospitalizations (**Figure 2A**) that occurs during the fall.

**Figure 2.**
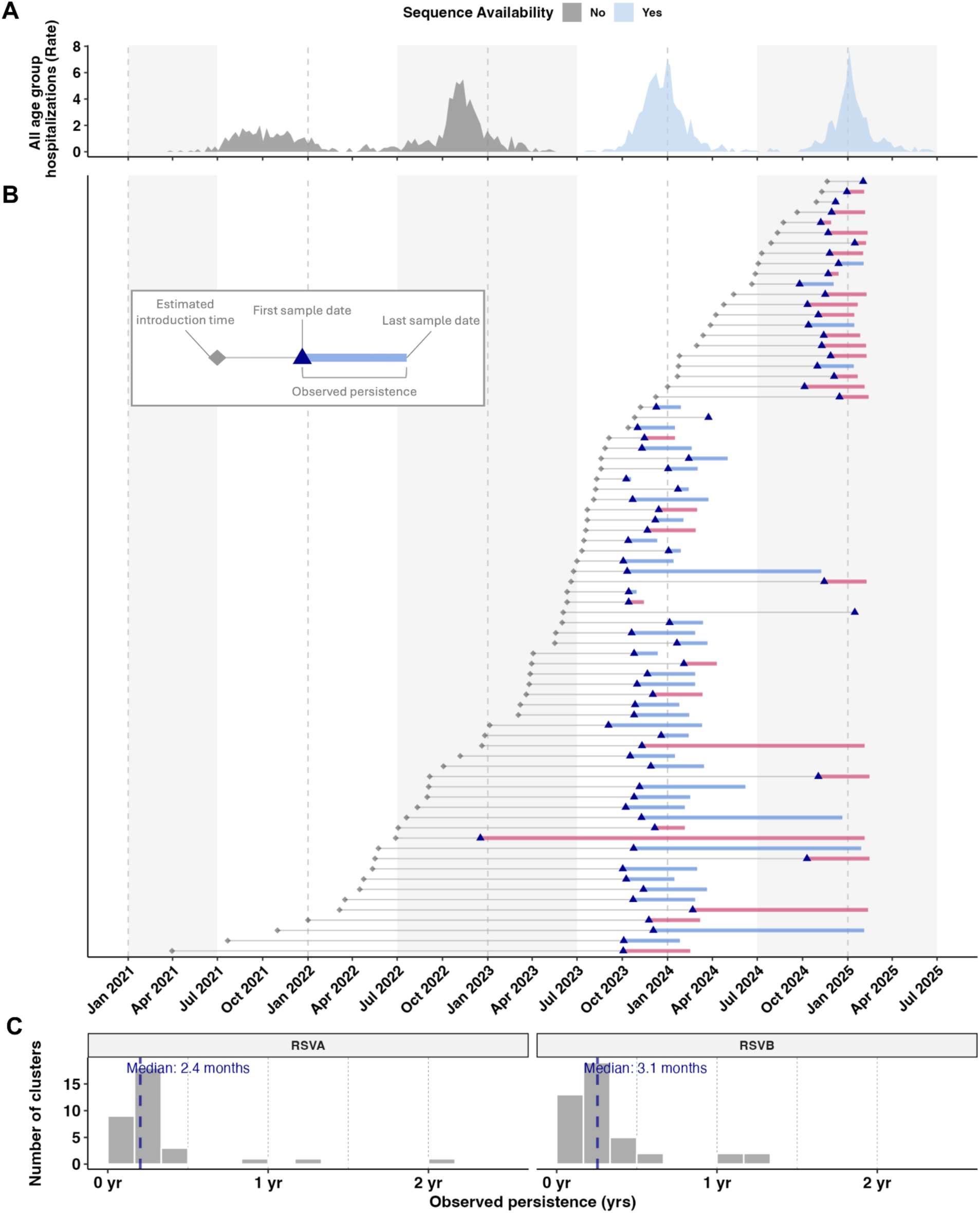
Introduction and observed persistence of Connecticut clades. (A) All age-group hospitalization rates in Connecticut from 2021 – 2025, based on data from RSV-NET and colored by sequence availability. (B) Summary of local Connecticut clades (RSV-B = 43, RSV-A = 33). Persistence bars are colored by subtype (RSV-A, pink; RSV-B, blue). Each vertical dotted line represents the start of a calendar year mark; RSV epidemic seasons are indicated by a light grey background. (C) Histogram of observed persistence duration; vertical dashed lines represent the median duration of lineage persistence.

We defined local lineage circulation by two different measures: observed (sampled) and estimated (phylogeographic) persistence. When measured by observed persistence, defined as the time between first and last sequenced case within a unique clade, we estimated median observed persistence to be around 2.4 months for RSV-A and 3.1 months for RSV-B in Connecticut (**Figure 2C**). However, the estimated persistence (time to the most recent common ancestor [tMRCA] of the lineage to last detection) was substantially longer than observed persistence, lasting about a year (median duration = 11.3 months RSV-A; 10.2 months RSV-B) (**Supplemental Figures 3 and 4**). While this suggests that most RSV lineages persist only during a single transmission season, our phylogeographic estimates suggest the inferred introductions of these lineages into Connecticut may have occurred 9-10 months prior to detection.

We quantified the monthly difference in the number of unique circulating lineages between RSV-A and RSV-B subtypes across two study seasons in Connecticut. In the first season, RSV-B dominated, its lineages outnumbering RSV-A until November 2024 (**Supplemental Figure 5A**). This difference peaked in December 2023, when we identified 25 unique RSV-B lineages compared to only 6 from RSV-A. Although the magnitude of this gap fluctuated throughout the year, RSV-B lineages continuously maintained higher lineage diversity than RSV-A. We observed a dramatic switch in December 2024, when observed circulating RSV-A lineages outnumbered RSV-B by a margin of 15. Using the phylogeographic estimates of lineage persistence (**Supplemental Figures 3 and 4**), we anticipate that this subtype dominance switch (RSV-B in 2023/24 to RSV-A in 2024/25) occurred eight months prior to when it was observed (**Supplemental Figure 5B**). We observed parity between the numbers of circulating lineages from both subtypes in March 2024; thereafter, RSV-A predominated and peaked in January 2025. This marked increase in introduced and circulating RSV-A lineages directly corresponds to the dominance of RSV-A within the sampled sequences for the season.

### Trans-seasonal persistence

We observed seven distinct clades, three RSV-A and four RSV-B, that we detected during both seasons of our surveillance period in Connecticut (**Figure 3**). Due to the fact that there are fewer sequences outside of Connecticut during our 2023-25 sampling period, some of the clades may be biased and not represent trans-seasonal persistence. A gap in the sequencing of RSV samples from important source locations can artificially extend our local clade tMRCA estimates to more ancestral nodes, whereas in reality lineages may leave Connecticut and return the following year. We noted three (non-mutually exclusive) general phylogenetic structures that provide some level of evidence for trans-seasonal persistence: (1) A relatively recent tMRCA estimated to have occurred no more than one season prior to the earliest sequence (clades A2, B3, and B4); (2) Recent sequences that extend from within, not basal to, the major cluster from the prior season (clades B1 and B3); (3) Clades with more than one sequence from both seasons (clade B2). These clades (A2 and B1-4) are thus more likely to represent true trans-seasonal persistence than clades with a relatively older tMRCA, extend through basal branches, and only one sequence sampled during one of the seasons (A1 and A3; **Figure 3**). Additionally, we found that the age distribution associated with these trans-seasonal persistent clades skewed towards older children and adults (**Figure 3**), with this pattern more prominent in RSV-B than RSV-A, and not toward the youngest age groups (under 1 or 12-35 months) as expected from hospitalization rates(*22*). Thus, additional research and increased nation-wide genomic surveillance is required to estimate the frequency of lineages that can persist between RSV transmission seasons.

**Figure 3.**
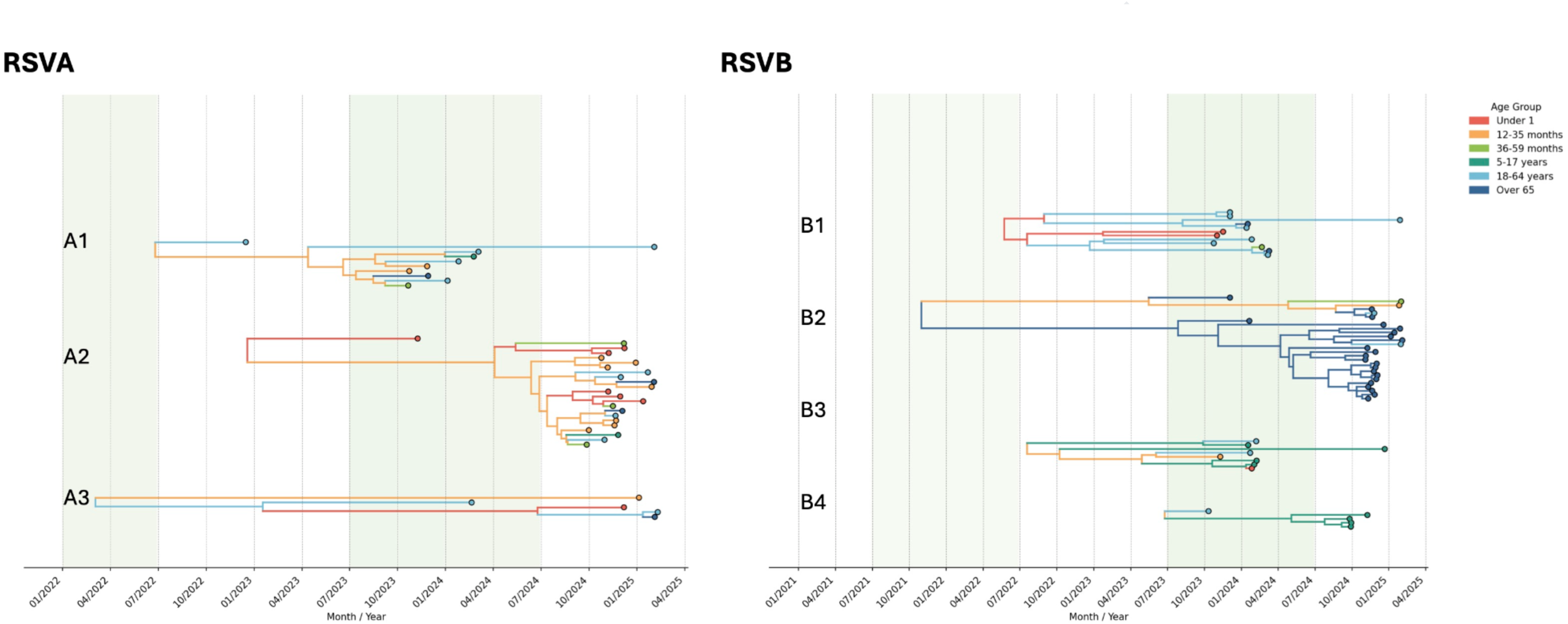
Trans-seasonal persistence of RSV clades. Maximum clade credibility (MCC) trees illustrating seven potential trans-seasonal persistent clades (three RSV-A and four RSV-B). Clades are defined by tips sampled across more than one RSV season from a single inferred introduction event. Tips are colored according to the age group from which the sequence was sampled. Internal branches are colored by the inferred ancestral age group, which is derived from the larger global phylogenetic tree.

### Key age group maintaining local transmission

To understand how RSV is maintained locally, we conducted an additional discrete trait phylodynamic analysis analysis, using age groups to quantify inter-group virus transmission events (Markov jumps) using all locally established lineages in Connecticut across the two RSV epidemic seasons. Across both RSV subtypes, the 12-35-month age group consistently emerged as the largest contributor to onward transmission to other age groups, with over 51% of all between-age-group viral transmission events originating from this age group (**Figure 4A**). The under-1-year group was the second leading source of transmission, accounting for 31-34% of transmission across both subtypes for all age groups.

**Figure 4.**
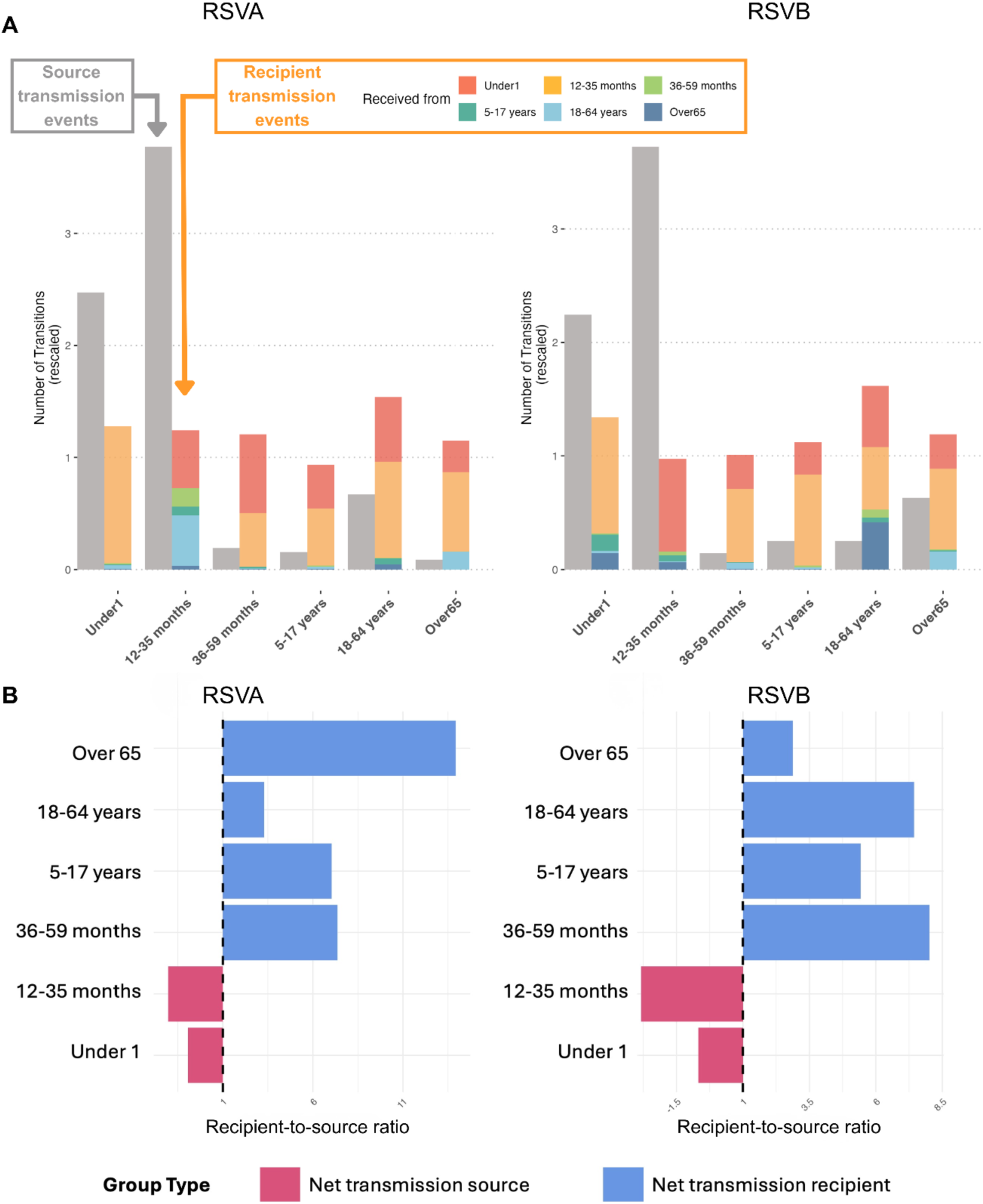
Inter-age-group virus transmission. (A) Age-stratified bar chart displaying the absolute number of virus transmission events between each age group estimated from the discrete phylodynamic analyses. Source transmission events are colored in grey and the recipient transmission events are colored by the source age-group. (B) Mirrored transmission ratio derived from panel (A), characterizing the age-specific role in transmission. We represent net recipients (ratio > 1) as the raw ratio and net sources (ratio < 1) as the negative inverse of the ratio. This transformation centers the data at 1.

While all age groups received comparable numbers of virus transmission events, their role as sources varied substantially. The recipient-to-source transmission ratio delineates distinct age-specific roles. The two youngest groups (35 months and younger) consistently functioned as net transmission sources, and the 36 months and older groups were net recipients (**Figure 4B**). This is because the older groups contribute minimally to onward transmission across age groups, functionally acting like ecological sinks. However, pairwise comparison among the older groups (36 months and older) showed higher variability in transmission directionality across age groups and subtypes (**Supplemental Figure 6**). While the recipient-to-source transmission ratios generally centered around 1, the low number of total transmission events in these cohorts made the observed directionality (net recipient vs net source) highly sensitive to fluctuations. We performed a phylogenetic “tip swap” sensitivity analysis to determine whether our findings reflect true transmission dynamics or are driven by age-group sampling proportions (**Table S1**). We generated a null model by randomly permuting age group labels across the tree tips while maintaining the original sampling distribution. Comparing this null distribution to our observed reconstructions isolates true transmission signals from those expected solely by sampling frequency. This analysis suggested the 12–35-month-olds would appear disproportionately responsible for transmission while the under-1-year group would contribute negligibly if we disregarded genetic clustering (**Supplemental Figure 7**). This provides some reassurance that patterns we infer are robust to possible sampling bias. Collectively, we show that integrating genomic data with non-genomic data in a phylodynamic framework allows us to investigate the diverse contributions of multiple age groups. Specifically, age groups under 1 and 18-65 years of age also play a pivotal role in propagating onward transmission throughout different sections of the population and shaping population-level RSV genetic diversity.

## Discussion

Using a hospital-based genomic surveillance system that we established to deeply sequence RSV across two seasons, we evaluated local RSV transmission dynamics and age-specific patterns through phylodynamic approaches. Our phylogenetic reconstructions reveal a critical decoupling of introduction and detection. RSV introductions into Connecticut occur year-round. However, we typically do not detect introduced clades until late fall or winter months, and detected clades, on average, persist for three months. Furthermore, across all sampled years, children aged 12-35 months emerged as the lead source of onward viral transmission between age groups, closely followed by infants under 1 year. These findings challenge the paradigm of strict seasonal RSV circulation, demonstrate the critical role of young children in shaping population-level RSV genetic diversity, and underscore the value of age-stratified genomic surveillance for informing control strategies. Ultimately, maintaining year-round genomic surveillance is essential to detect emerging clades early and to capture the full landscape of viral introduction and persistence.

Our findings also provide critical context for broader U.S. spatiotemporal trends of RSV. Current understanding of the spatially structured timing of U.S. RSV epidemics posits two primary hypotheses: propagating transmission waves originating in the southeast, or local initiation of seasonal outbreaks governed by factors such as climate or susceptible population dynamics(*14*, *23*, *24*). Our genomic analyses provide further insight into this unresolved question. We demonstrate strong evidence that the local U.S. RSV epidemic cycle relies on frequent, year-round, independent introduction events often stemming from multiple different U.S. regional sources, a pattern that case data alone cannot distinguish. Collectively, our findings weigh against a single explosive expansion via a “spatial wave” originating from the Southeast and progressing to the Northeast through the Mid-Atlantic region.

We decouple the continuous RSV introduction events from clinical detection of the virus, and present that the typical RSV epidemic season reflects the timing of symptomatic presentation, not the initial seeding of the virus. The usual fall/winter seasonal emergence likely occurs when the virus successfully establishes high-level circulation within populations most susceptible to disease, such as young children and older adults, potentially catalyzed by local, environmental, or host-susceptibility factors. Our data indicate low-level cryptic circulation outside of the typical epidemic window, a hypothesis supported by evidence of asymptomatic shedding in non-infant contacts in existing household-based studies(*25*, *26*), and that trans-seasonal clades were present in a broad age range, rather than strictly skewing toward the highest-risk populations. Substantially reduced testing efforts in this period further contribute to the low probability of detection in the summer.

Our analysis revealed that importation events with limited persistence of less than a year drive the majority (90%) of observed clades, which aligns with existing evidence from different countries, including Australia(*27*), Kenya(*28*), and a multi-country study(*29*). Conversely, a small but notable fraction (11%) demonstrated local trans-seasonal persistence. This observation is not unprecedented, having been reported in South Africa(*29*). However, its interpretation requires careful consideration, as this persistence may either represent genuine local, sustained transmission or be an artifact of limited background sequence availability. Specifically, the underrepresentation may mask an “out-and-in” event, where two (or more) separate introduction events appear as continuous local persistence because of a sampling gap in the location in which it was maintained. While correcting for this possible underrepresentation would yield more precise clade-specific parameter estimates (e.g., number, duration, and timing of introduction), the overall interpretation of our key research questions is not fundamentally altered; this is because Bayesian phylogenetic inference methods ensured robust inferences by pooling information across clades in addition to clade-specific parameters(*30*, *31*). However, the precise mechanism of persistence holds relevance for understanding broader surveillance implications in the local setting; filling this knowledge gap will require expanded nation-wide surveillance.

We demonstrate that toddlers (12-35 months) significantly shape circulating RSV genetic diversity and are central to forward transmission. While previous studies primarily explored age-group-specific roles in household cohorts, our population-level analysis expands on these findings. Consistent with prior household transmission studies highlighting the role of young children in household spread to adults(*32*) and older children’s (2-13 years) contribution to infant infections(*33*), we further characterize age group-specific transmission roles and their influence on population-level RSV genetic diversity. Critically, incorporating genetic clustering unmasks a non-negligible transmission contribution from infants (under 1 year) to all age groups and highlights the role of the adult (18-64 years) age group in onward spread, particularly to individuals over 65. The observed transmission directionality from the youngest age groups to older populations may account for the approximately one-month lag in the peak of RSV cases in the under-5 age group versus older adults(*17*). Our detailed characterization of age-specific connectivity and directionality provides a crucial foundation for tailoring future RSV immunization strategies.

There are additional limitations to our study. Primarily, the absence of standardized genomic surveillance data across U.S. states impedes our ability to delineate the exact origin or granular scale relatedness across states or regions most closely related to Connecticut in RSV transmission. Generalizability is also limited. We cannot confirm whether these local transmission dynamics represent broader U.S. contexts, temperate regions, or how patterns shift in distinct settings, such as tropical climates. Future investigations must characterize observed patterns across diverse geographical and climatic regions. Furthermore, the two-year duration of surveillance data leaves the subtle yet potentially critical differences in transmission dynamics driven by RSV-A versus RSV-B subtypes and how these relate to different age groups uncharacterized. In contrast, studies of influenza using longitudinal data have shown that certain subtypes are associated with younger ages of infection and smaller epidemics(*20*). Extended surveillance, incorporating subtype characterization, is necessary to better understand how changes in subtype-specific immunity influence the seasonal RSV epidemic.

Our findings emphasize that toddlers, alongside infants, primarily drive local genetic diversity and spread. This justifies prevention strategies that expand beyond reducing disease burden to actively prioritizing reducing transmission and infection, although current immunization approaches effectively reduce RSV disease severity in infants(*34*) and older adults(*35*). Furthermore, as immunization expands, there is the potential for an increase in immunologic pressure(*29*); Proactive monitoring from public health systems to track RSV introduction and circulation would enable timely intervention against potentially resistant clades before they become clinically widespread. Understanding local-level clade persistence is vital for determining whether novel resistant strains become locally established or contribute to regional spread. Ultimately, we establish that solidifying local-level RSV genomic surveillance is a critical tool for enhancing effective long-term control of RSV.

## Materials and Methods

### Ethics statement

The study protocol for pathogen genomic sequencing of remnant diagnostic samples was reviewed and approved by the Yale University institutional review board (IRB). All specimens were de-identified, remnant specimens, used previously for diagnostic testing or IRB-approved human subjects research, in accordance with Yale University IRB-exempt protocol #2000033281.

### Sample collection and RNA extraction

Clinical samples were obtained from the Yale New Haven Hospital (YNNH) between October 2023 and March 2025, corresponding with the peak of RSV season. YNNH performs diagnostic nucleic acid amplification tests for hospitals across the Yale New Haven Health System, covering most of southern Connecticut. Nasopharyngeal swabs were flagged for study if they were confirmed positive for RSV during routine diagnostic testing. Remnant samples were stored in viral transport media (BD Universal Viral Transport Medium) and transported to the Yale School of Public Health, where they were stored at −80°C until extraction for genomic sequencing.

Samples were defrosted at 4°C overnight and RNA was extracted using 300 μL of each sample on the Kingfisher Flex 96 automated extraction instrument (ThermoFisher Scientific, Waltham, MA) with either the MagMax Viral/Pathogen Total Nucleic Acid Kit (ThermoFisher, product code A42352) with no modifications to the manufacturers. protocol or the Monarch Mag Viral DNA/RNA Extraction Kit (New England Biolabs, product code: T4010), omitting the addition of carrier RNA as written in the manufacturer’s protocol, and eluted in 75μL of elution buffer.

### Multiplexed primer design for amplicon sequencing

Our custom primer set was modified from the Maloney protocol(*36*) primer set to improve the consistency of coverage of the RSV G gene across genetically divergent lineages. We tested our primer set with commercial RSV-A (ATCC VR-1540) / RSV-B (ATCC VR-3381) standards, both individually and as a multiplex, and then validated with 182 mixed-genotype (A and B) clinical samples to confirm that we could sequence without the need for prior subtyping. Our primer sequences are available in **Supplementary Data 1 (RSV-A)** and **Supplementary Data 2 (RSV-B)**).

### DNA library preparation and amplicon sequencing

DNA libraries were prepared using the Illumina COVIDSeq DNA prep kit (Illumina, San Diego, CA) with protocol modifications as previously described(*37*).

Briefly, template RNA was synthesized into cDNA and subsequently amplified in two separate multiplex PCR reactions, with each primer pool containing non-overlapping sections of the tiled primer scheme. After amplification, PCR products were combined in equal parts and subjected to a 3-minute tagmentation. Tagmented amplicons were purified using beads provided in the CovidSeq Kit, followed by library amplification and adapter ligation with IDT for Illumina Unique Dual Indexes (Illumina, San Diego, CA). Individual indexed libraries were pooled in equal proportions and underwent a small fragment removal, selecting for DNA fragments greater than 300bp. The purified library pool was quantified using a 1x dsDNA High-Sensitivity Assay kit (Applied Biosystems, Carlsbad, CA) on the Qubit 4 Fluorometer (Thermo Fisher, Waltham, MA), and fragment distribution was verified using a dsDNA High-Sensitivity kit on the 2100 Agilent Bioanalyzer Instrument (Agilent, Santa Clara, CA). Pooled libraries were sequenced in 150 bp paired-end reads at the Yale Center for Genome Analysis on an Illumina NovaSeq (Illumina, San Diego, CA), with an average of 1 million reads generated per library.

### Consensus sequence generation and calling

After preliminary subtype identification with MASH(*38*) (minhash match probability <= 1e-50), reads were aligned to the appropriate reference (RSV-A: KY654518.1; RSV-B: OP975389.1) using BWA-MEM (v.2.2.1)(*39*) and SAMtools (v1.15.1)(*40*). Read sets that ambiguously matched to both references were aligned to both. Amplicon sequencing data were filtered (using defaults; Q>20 over a sliding window of 4, minimum read length of 50% of the average length). After trimming of primer sequences using iVar (v.1.4.2)(*41*), variants were called and filtered (Phred score Q>10 and read depth >10) using BCFtools (v.1.21)(*42*). Viral consensus genomes and intra-host variants were constructed using iVar (v.1.4.2)(*41*). Read depth, coverage, and summary statistics were calculated using SAMtools(*40*). Further data analyses and visualizations were carried out using the tidyverse suite (v.2.0.0)(*43*). For ambiguously subtyped samples, alignments with < 80% coverage were discarded and primary and secondary alignments compared; samples with secondary coverage of >=95% the primary were determined to be potential co-infections. The alignment and calling pipeline was constructed using Snakemake(*44*) and Singularity(*45*) and is available from https://github.com/sethnr/YSPH_RSV/.

### Initial processing of genomic data

We downloaded all RSV whole-genome sequences with at least 90% coverage, reported country, and year of sampling from GenBank on 11 April 2025. We combined this dataset with our newly generated 910 sequences and aligned them to reference genomes; references were updated to PP109421.1 (RSV-A) and OP975389.1 (RSV-B) for consistency with assigned references on Pathoplexus. Alignment was conducted using MAFFT v7.505(*46*), and then we used AliView(*47*) to retain only the coding regions and manually verified that each gene was accurately translated to the corresponding amino acid. This yielded a final alignment length of 13,680 bases (RSV-A) and 13,719 bases (RSV-B), and a dataset size of 7746 RSV-A and 6227 RSV-B sequences.

We constructed a maximum likelihood tree using IQ-TREE v2.1.2(*48*), employing a general time reversible substitution model with gamma-distributed rate variation (GTR- 𝛤) in alignment with a previous multi-country RSV study(*29*). We used the resulting tree to assess the temporal signal in TempEst(*49*), and removed sequences identified as molecular clock outliers(*50*).

### Epidemiological data and down-sampling

We categorized sequences into eight location categories to investigate RSV local dynamics and lineage establishment in Connecticut. Our primary category, U.S. Connecticut, included all available Connecticut sequences that met the inclusion criteria described in the section above. Those remaining U.S. sequences were categorized into five regions to minimize sparsity: Northeast, Midwest, West, Southeast, and Southwest (**Supplemental Figure 8**). Non-U.S. sequences were broadly classified into northern and southern hemisphere sequences to account for epidemiological differences between hemispheres.

To contextualize Connecticut lineages in relation to other sequences, we downsampled background sequences in the dataset by location and RSV season to balance representation across geographic categories to mitigate sampling bias. All available Connecticut sequences, including those generated in this study and from Lauring et al.(*51*), were exempted from downsampling. For each non-Connecticut U.S. region, we randomly selected up to 70 sequences per season. For each hemisphere-level category, we randomly selected up to 100 sequences per season. We defined the RSV season by hemisphere for sequences with a known collection month. For northern hemisphere countries (including the U.S.), a season was defined as July 1^st^ of one year to June 30^th^ of the next. For the southern hemisphere, a season was defined by the calendar year. In the case a season had fewer sequences than the specified cap, all available sequences were retained. We included sequences for which only the collection year was known to supplement the downsampled dataset to increase overall temporal resolution. This downsampling resulted in 4119 genomes for RSV-A and 3597 for RSV-B (**Tables S2-S4**), including 894 total sequences from Connecticut.

We obtained age-stratified RSV testing rates in Connecticut from January 2023 to July 2025 via the PopHIVE data hub(*52*). We also utilized data from the population-based RSV hospitalization surveillance network (RSV-NET)(*53*) to characterize all-age-group hospitalization rates in Connecticut (January 2021 – July 2025).

### Phylogenetic analysis

We estimated time-calibrated evolutionary histories using the Bayesian Evolutionary Analysis by Sampling Trees (BEAST X)(*30*) platform, along with the high-performance BEAGLE 4.0.1(*54*). For tree inference, we applied a GTR-𝛤 nucleotide substitution model, a Hamiltonian Monte Carlo relaxed molecular clock model(*55*), and a non-parametric Skygrid coalescent tree prior(*56*) for phylogeny across both RSV-A and RSV-B. To reflect the seasonality, we allowed the skygrid to change twice per year. We used tip-date sampling for sequences with only the year of collection. For each RSV subtype, we ran two independent chains of at least 100 million iterations We discarded the initial ∼10% of each chain as burn-in after manual assessment of mixing and convergence via Tracer 1.7(*57*), and combined the resulting chains using LogCombiner. We used TreeAnnotator to generate a maximum clade credibility (MCC) tree.

### Identifying unique Connecticut lineages: phylogeographic inference

To reconstruct the importation dynamics of RSV into Connecticut, we performed an asymmetric discrete trait analysis (DTA) model implemented in BEAST X. We used 1000 random trees from the posterior distribution of the above analysis as the empirical tree distribution for this model to improve computational efficiency while still incorporating phylogenetic uncertainty. We assigned taxa into eight geographical categories as discussed above: Connecticut, five U.S. regions, and two hemisphere categories (non-U.S. northern hemisphere and southern hemisphere). We ran two independent chains with 5 million iterations each. Convergence was assessed using Tracer, chains were combined using LogCombiner, and the MCC tree summary was generated using TreeAnnotator. We used all packages distributed as part of BEAST X(*30*).

We defined Connecticut clades as monophyletic clades of at least three sequences, all from Connecticut, where the ancestry of the entire cluster remained continuously within Connecticut. We defined the introduction time as the height of the earliest node inferred to be in Connecticut, where the preceding node was any other location. We summarized transitions between states using the TreeMarkovJumpHistoryAnalyzer(*58*).

### Inter-age-group dynamics

We conducted a joint DTA analysis on each Connecticut clade together to analyze transmission dynamics by age group. We assigned each sequence to one of six age groups: under 1 year (0–11 months), 12–35 months, 36–59 months, 5–17 years, 18–64 years, and over 65 years. We employed the same general configuration and workflow used for the phylogeographic DTA.

### Data and code availability

New RSV sequences have been deposited in GenBank via Pathoplexus and are publicly available (https://doi.org/10.62599/PP_SS_206.2). We used the R statistical software (v4.4.1) and Python for all statistical analyses and visualization. Data and code used in this study are publicly available on GitHub: https://github.com/jiyekwon/Yale_RSV_age.

## Acknowledgments

We would like to thank Adam Lauring, Leigh Papalambros, Basmah Safdar, and Wesley Self for sharing state-level location information for the sequences used in JAMA research letter “Genomic Characterization of RSV in the US by Vaccination Status”.

This publication was made possible by the New England Pathogen Genomics Center of Excellence (U.S. CDC NU50CK000629) and the Office of Advanced Molecular Detection, Centers for Disease Control and Prevention through Cooperative Agreement Number CK22-2204. This project was additionally supported by the Merck Investigator Studies Program contracts 103595 (NDG, VEP, DMW) and 103622 (VEP, DMW, NDG), and UMC Utrecht Strategic Network Development Grant D-24-700098 (JK, LB), as well as the National Institutes of Health/ National Institute of Allergy and Infectious Diseases grants R01AI137093 (DMW, VEP), R01AI179874 (CRO, DMW, NDG), and 1S10OD030363-01A1. The funders had no role in study design, data collection and analysis, decision to publish, or preparation of the manuscript. The findings and conclusions in this report are those of the author(s) and do not necessarily represent the official position of the Centers for Disease Control and Prevention or the National Institutes of Health.

## Author contributions

Conceptualization: JK, VEP, DMW, NDG, VH, SR

Methodology: JK, KL, PL, VEP, DMW, NDG, VH

Resources: VEP, DMW, LB

Data Curation: ED, JK, MB, KL, DF, WS, CRO, SR

Writing – original draft: JK

Writing – review & editing: All authors.

Visualization: JK, PL, VH

Supervision: PL, LB, VH, VEP, DMW, NDG

Project administration: SR

Funding acquisition: VEP, DMW, NDG

## References

1. Y. Li, X. Wang, D. M. Blau, M. T. Caballero, D. R. Feikin, C. J. Gill, S. A. Madhi, S. B. Omer, E. A. F. Simões, H. Campbell, A. B. Pariente, D. Bardach, Q. Bassat, J.-S. Casalegno, G. Chakhunashvili, N. Crawford, D. Danilenko, L. A. H. Do, M. Echavarria, A. Gentile, A. Gordon, T. Heikkinen, Q. S. Huang, S. Jullien, A. Krishnan, E. L. Lopez, J. Markić, A. Mira-Iglesias, H. C. Moore, J. Moyes, L. Mwananyanda, D. J. Nokes, F. Noordeen, E. Obodai, N. Palani, C. Romero, V. Salimi, A. Satav, E. Seo, Z. Shchomak, R. Singleton, K. Stolyarov, S. K. Stoszek, A. von Gottberg, D. Wurzel, L.-M. Yoshida, C. F. Yung, H. J. Zar, M. Abram, J. Aerssens, A. Alafaci, A. Balmaseda, T. Bandeira, I. Barr, E. Batinović, P. Beutels, J. Bhiman, C. C. Blyth, L. Bont, S. S. Bressler, C. Cohen, R. Cohen, A.-M. Costa, R. Crow, A. Daley, D.-A. Dang, C. Demont, C. Desnoyers, J. Díez-Domingo, M. Divarathna, M. du Plessis, M. Edgoose, F. M. Ferolla, T. K. Fischer, A. Gebremedhin, C. Giaquinto, Y. Gillet, R. Hernandez, C. Horvat, E. Javouhey, I. Karseladze, J. Kubale, R. Kumar, B. Lina, F. Lucion, R. MacGinty, F. Martinon-Torres, A. McMinn, A. Meijer, P. Milić, A. Morel, K. Mulholland, T. Mungun, N. Murunga, C. Newbern, M. P. Nicol, J. K. Odoom, P. Openshaw, D. Ploin, F. P. Polack, A. J. Pollard, N. Prasad, J. Puig-Barberà, J. Reiche, N. Reyes, B. Rizkalla, S. Satao, T. Shi, S. Sistla, M. Snape, Y. Song, G. Soto, F. Tavakoli, M. Toizumi, N. Tsedenbal, M. van den Berge, C. Vernhes, C. von Mollendorf, S. Walaza, G. Walker, H. Nair, Global, regional, and national disease burden estimates of acute lower respiratory infections due to respiratory syncytial virus in children younger than 5 years in 2019: a systematic analysis. Lancet 399, 2047–2064 (2022).

2. F. P. Havers, M. Whitaker, M. Melgar, H. Pham, S. J. Chai, E. Austin, J. Meek, K. P. Openo, P. A. Ryan, C. Brown, K. Como-Sabetti, D. M. Sosin, G. Barney, B. L. Tesini, M. Sutton, H. K. Talbot, R. Chatelain, P. Daily Kirley, I. Armistead, K. Yousey-Hindes, M. L. Monroe, V. Tellez Nunez, R. Lynfield, C. L. Esquibel, K. Engesser, K. Popham, A. Novak, W. Schaffner, T. M. Markus, A. Swain, M. E. Patton, L. Kim, Burden of Respiratory Syncytial Virus–Associated Hospitalizations in US Adults, October 2016 to September 2023. JAMA Netw. Open 7, e2444756–e2444756 (2024).

3. H. H. Nam, M. G. Ison, Respiratory syncytial virus infection in adults. BMJ 366, l5021 (2019).

4. H. J. Zar, F. Cacho, T. Kootbodien, A. Mejias, J. R. Ortiz, R. T. Stein, T. V. Hartert, Early-life respiratory syncytial virus disease and long-term respiratory health. Lancet Respir. Med. 12, 810–821 (2024).

5. “Respiratory syncytial virus” in Red Book: 2024–2027 Report of the Committee on Infectious Diseases (American Academy of Pediatrics 345 Park Blvd, Itasca, IL 60143, 2024), pp. 713–721.

6. K. E. Fleming-Dutra, J. M. Jones, L. E. Roper, M. M. Prill, I. R. Ortega-Sanchez, D. L. Moulia, M. Wallace, M. Godfrey, K. R. Broder, N. K. Tepper, O. Brooks, P. J. Sánchez, C. N. Kotton, B. E. Mahon, S. S. Long, M. L. McMorrow, Use of the Pfizer Respiratory Syncytial Virus Vaccine During Pregnancy for the Prevention of Respiratory Syncytial Virus-Associated Lower Respiratory Tract Disease in Infants: Recommendations of the Advisory Committee on Immunization Practices - United States, 2023. MMWR Morb. Mortal. Wkly. Rep. 72, 1115–1122 (2023).

7. Melgar M, Britton A, Roper LE, Talbot HK, Long SS, Kotton CN, Havers FP, Use of Respiratory Syncytial Virus Vaccines in Older Adults: Recommendations of the Advisory Committee on Immunization Practices — United States, 2023. MMWR Morb. Mortal. Wkly. Rep. 72, 793–801 (2023).

8. D. L. Moulia, R. Link-Gelles, H. Y. Chu, D. Jamieson, O. Brooks, S. Meyer, E. S. Weintraub, D. K. Shay, M. M. Prill, E. S. Thomas, D. Hutton, I. R. Ortega-Sanchez, A. MacNeil, M. L. McMorrow, J. M. Jones, Use of Clesrovimab for Prevention of Severe Respiratory Syncytial Virus-Associated Lower Respiratory Tract Infections in Infants: Recommendations of the Advisory Committee on Immunization Practices - United States, 2025. MMWR Morb. Mortal. Wkly. Rep. 74, 508–514 (2025).

9. A. Britton, L. E. Roper, C. N. Kotton, D. W. Hutton, K. E. Fleming-Dutra, M. Godfrey, I. R. Ortega-Sanchez, K. R. Broder, H. K. Talbot, S. S. Long, F. P. Havers, M. Melgar, Use of Respiratory Syncytial Virus Vaccines in Adults Aged ≥60 Years: Updated Recommendations of the Advisory Committee on Immunization Practices - United States, 2024. MMWR Morb. Mortal. Wkly. Rep. 73, 696–702 (2024).

10. J. M. Jones, K. E. Fleming-Dutra, M. M. Prill, L. E. Roper, O. Brooks, P. J. Sánchez, C. N. Kotton, B. E. Mahon, S. Meyer, S. S. Long, M. L. McMorrow, Use of Nirsevimab for the Prevention of Respiratory Syncytial Virus Disease Among Infants and Young Children: Recommendations of the Advisory Committee on Immunization Practices - United States, 2023. MMWR Morb. Mortal. Wkly. Rep. 72, 920–925 (2023).

11. P. K. Munywoki, D. C. Koech, C. N. Agoti, C. Lewa, P. A. Cane, G. F. Medley, D. J. Nokes, The source of respiratory syncytial virus infection in infants: a household cohort study in rural Kenya. J. Infect. Dis. 209, 1685–1692 (2014).

12. C. N. Agoti, M. V. T. Phan, P. K. Munywoki, G. Githinji, G. F. Medley, P. A. Cane, P. Kellam, M. Cotten, D. J. Nokes, Genomic analysis of respiratory syncytial virus infections in households and utility in inferring who infects the infant. Sci. Rep. 9, 10076 (2019).

13. Z. Zheng, J. L. Warren, I. Artin, V. E. Pitzer, D. M. Weinberger, Relative timing of respiratory syncytial virus epidemics in summer 2021 across the United States was similar to a typical winter season. *Influenza Other Respi*. Viruses 16, 617–620 (2022).

14. V. E. Pitzer, C. Viboud, W. J. Alonso, T. Wilcox, C. J. Metcalf, C. A. Steiner, A. K. Haynes, B. T. Grenfell, Environmental Drivers of the Spatiotemporal Dynamics of Respiratory Syncytial Virus in the United States. PLoS Pathog. 11, e1004591 (2015).

15. C. A. Panozzo, A. L. Fowlkes, L. J. Anderson, Variation in Timing of Respiratory Syncytial Virus Outbreaks: Lessons From National Surveillance. Pediatr. Infect. Dis. J. 26, S41–S45 (2007).

16. D. M. Weinberger, J. L. Warren, C. A. Steiner, V. Charu, C. Viboud, V. E. Pitzer, Reduced-Dose Schedule of Prophylaxis Based on Local Data Provides Near-Optimal Protection Against Respiratory Syncytial Virus. Clin. Infect. Dis. 61, 506–514 (2015).

17. K. Li, V. E. Pitzer, D. M. Weinberger, Exploring RSV Transmission Patterns in Different Age Groups in the United States. J. Infect. Dis. 232, 700–708 (2025).

18. V. Hill, S. Dellicour, M. Giovanetti, N. D. Grubaugh, Phylogenetic insights into the transmission dynamics of arthropod-borne viruses. Nat. Rev. Genet. 27, 47–61 (2026).

19. P. Lemey, N. Ruktanonchai, S. L. Hong, V. Colizza, C. Poletto, F. Van den Broeck, M. S. Gill, X. Ji, A. Levasseur, B. B. Oude Munnink, M. Koopmans, A. Sadilek, S. Lai, A. J. Tatem, G. Baele, M. A. Suchard, S. Dellicour, Untangling introductions and persistence in COVID-19 resurgence in Europe. Nature 595, 713–717 (2021).

20. T. Bedford, S. Riley, I. G. Barr, S. Broor, M. Chadha, N. J. Cox, R. S. Daniels, C. P. Gunasekaran, A. C. Hurt, A. Kelso, A. Klimov, N. S. Lewis, X. Li, J. W. McCauley, T. Odagiri, V. Potdar, A. Rambaut, Y. Shu, E. Skepner, D. J. Smith, M. A. Suchard, M. Tashiro, D. Wang, X. Xu, P. Lemey, C. A. Russell, Global circulation patterns of seasonal influenza viruses vary with antigenic drift. Nature 523, 217–220 (2015).

21. M. A. Mufson, C. Orvell, B. Rafnar, E. Norrby, Two distinct subtypes of human respiratory syncytial virus. J. Gen. Virol. 66 (Pt 10), 2111–2124 (1985).

22. Yale School of Public Health, Infectious Diseases, PopHIVE (2025). https://www.pophive.org/infectious-diseases/respiratory-syncytial-virus#viral-activity-by-age.

23. Z. Zheng, V. E. Pitzer, E. D. Shapiro, L. J. Bont, D. M. Weinberger, Estimation of the Timing and Intensity of Reemergence of Respiratory Syncytial Virus Following the COVID-19 Pandemic in the US. *JAMA Netw*. Open 4, e2141779 (2021).

24. R. E. Baker, A. S. Mahmud, C. E. Wagner, W. Yang, V. E. Pitzer, C. Viboud, G. A. Vecchi, C. J. E. Metcalf, B. T. Grenfell, Epidemic dynamics of respiratory syncytial virus in current and future climates. Nat. Commun. 10, 5512 (2019).

25. P. K. Munywoki, D. C. Koech, C. N. Agoti, N. Kibirige, J. Kipkoech, P. A. Cane, G. F. Medley, D. J. Nokes, Influence of age, severity of infection, and co-infection on the duration of respiratory syncytial virus (RSV) shedding. Epidemiol. Infect. 143, 804–812 (2015).

26. H. Otomaru, J. B. T. Sornillo, T. Kamigaki, S. L. P. Bado, M. Okamoto, M. Saito-Obata, M. T. Inobaya, E. Segubre-Mercado, P. P. Alday, M. Saito, V. L. Tallo, B. P. Quiambao, H. Oshitani, A. R. Cook, Risk of Transmission and Viral Shedding From the Time of Infection for Respiratory Syncytial Virus in Households. Am. J. Epidemiol. 190, 2536–2543 (2021).

27. M. Robertson, J. S. Eden, A. Levy, I. Carter, R. L. Tulloch, E. J. Cutmore, B. A. Horsburgh, C. T. Sikazwe, D. E. Dwyer, D. W. Smith, J. Kok, The spatial-temporal dynamics of respiratory syncytial virus infections across the east-west coasts of Australia during 2016-17. Virus Evol. 7, veab068 (2021).

28. N. Agoti Charles, R. Otieno James, M. Ngama, G. Mwihuri Alexander, F. Medley Graham, A. Cane Patricia, D. J. Nokes, Successive Respiratory Syncytial Virus Epidemics in Local Populations Arise from Multiple Variant Introductions, Providing Insights into Virus Persistence. J. Virol. 89, 11630–11642 (2015).

29. A. C. Langedijk, B. Vrancken, R. J. Lebbink, D. Wilkins, E. J. Kelly, E. Baraldi, A. H. Mascareñas de Los Santos, D. M. Danilenko, E. H. Choi, M. A. Palomino, H. Chi, C. Keller, R. Cohen, J. Papenburg, J. Pernica, A. Greenough, P. Richmond, F. Martinón-Torres, T. Heikkinen, R. T. Stein, M. Hosoya, M. C. Nunes, C. Verwey, A. Evers, L. Kragten-Tabatabaie, M. A. Suchard, S. L. Kosakovsky Pond, C. Poletto, V. Colizza, P. Lemey, L. J. Bont, E. Priante, K. Komissarova, K. W. Yun, P. Clement, M. Bauck, A. Gupta, U. Wadia, I. Rivero-Calle, M. Lumertz, K. Hasimoto, S. A. Madhi, on behalf of the, Inform- R. S. V. Study Group, The genomic evolutionary dynamics and global circulation patterns of respiratory syncytial virus. Nat. Commun. 15, 3083 (2024).

30. G. Baele, X. Ji, G. W. Hassler, J. T. McCrone, Y. Shao, Z. Zhang, A. J. Holbrook, P. Lemey, A. J. Drummond, A. Rambaut, M. A. Suchard, BEAST X for Bayesian phylogenetic, phylogeographic and phylodynamic inference. Nat. Methods 22, 1653–1656 (2025).

31. P. Lemey, A. Rambaut, A. J. Drummond, M. A. Suchard, Bayesian Phylogeography Finds Its Roots. PLoS Comput. Biol. 5, e1000520 (2009).

32. S. N. Cox, P. Roychoudhury, C. Frivold, Z. Acker, T. M. Babu, C. L. Boisvert, M. Carone, B. Ehmen, J. A. Englund, L. R. Feldstein, L. Gamboa, S. Grindstaff, H. M. Grioni, P. D. Han, K. L. Hoffman, H. G. Kim, J. L. Kuntz, N. K. Lo, C. M. Lockwood, K. McCaffrey, R. A. Mularski, T. L. Hatchie, S. L. Reich, M. A. Schmidt, N. Smith, L. M. Starita, A. Varga, N. Yetz, A. L. Naleway, A. A. Weil, H. Y. Chu, Household transmission and genomic diversity of respiratory syncytial virus (RSV) in the United States, 2022-2023. Clin. Infect. Dis., doi: 10.1093/cid/ciaf048 (2025).

33. I. K. Kombe, C. N. Agoti, P. K. Munywoki, M. Baguelin, D. J. Nokes, G. F. Medley, Integrating epidemiological and genetic data with different sampling intensities into a dynamic model of respiratory syncytial virus transmission. Sci. Rep. 11, 1463 (2021).

34. H. Xu, C. Aparicio, A. Wats, B. L. Araujo, V. E. Pitzer, J. L. Warren, E. D. Shapiro, L. M. Niccolai, D. M. Weinberger, C. R. Oliveira, Estimated effectiveness of nirsevimab against respiratory syncytial virus. *JAMA Netw*. Open 8, e250380 (2025).

35. D. Surie, W. H. Self, Y. Zhu, K. A. Yuengling, C. A. Johnson, C. G. Grijalva, F. S. Dawood, Investigating Respiratory Viruses in the Acutely Ill (IVY) Network, RSV vaccine effectiveness against hospitalization among US adults 60 years and older. JAMA 332, 1105–1107 (2024).

36. D. M. Maloney, G. Fernandes, S. Jasim, T. Williams, S. Namugenyi, M. Carr, J. R. Meyer, A. Sharma, L. Marshal, B. E. Nunley, H. Xie, M. Carvajal, H. D. Kunerth, A. Tanmoy, Á. O’Toole, A. Weixler, K. R. Lanter, G. Gonzalez, J. Sereewit, R. Guevara, M. Loose, P. Fanning, D. Benítez, J. C. Fernandez, P. Cárdenas, D. Hare, A. Greninger, T. Williams, G. Nebbia, A. C. Fries, P. McClure, P. Roychoudhury, X. Wang, S. Saha, R. Dewar, K. E. Templeton, A. Rambaut, ARTIC RSV amplicon sequencing reveals global RSV genotype dynamics. Wellcome Open Res. 10, 323 (2025).

37. N. F. G. Chen, L. Gagne, M. Doucette, S. Smole, E. Buzby, J. Hall, S. Ash, R. Harrington, S. Cofsky, S. Clancy, C. J. Kapsak, J. Sevinsky, K. Libuit, C. Chaguza, N. D. Grubaugh, D. J. Park, G. E. Gallagher, C. B. F. Vogels, Monkeypox virus multiplexed PCR amplicon sequencing (PrimalSeq). protocols.io, doi: 10.17504/protocols.io.5qpvob1nbl4o/v4 (2022).

38. B. D. Ondov, T. J. Treangen, P. Melsted, A. B. Mallonee, N. H. Bergman, S. Koren, A. M. Phillippy, Mash: fast genome and metagenome distance estimation using MinHash. Genome Biol. 17, 1–14 (2016).

39. H. Li, Aligning sequence reads, clone sequences and assembly contigs with BWA-MEM. arXiv [q-bio.GN] (2013).

40. H. Li, B. Handsaker, A. Wysoker, T. Fennell, J. Ruan, N. Homer, G. Marth, G. Abecasis, R. Durbin, The Sequence Alignment/Map format and SAMtools. Bioinformatics 25, 2078–2079 (2009).

41. N. D. Grubaugh, K. Gangavarapu, J. Quick, N. L. Matteson, J. G. De Jesus, B. J. Main, A. L. Tan, L. M. Paul, D. E. Brackney, S. Grewal, N. Gurfield, K. K. A. Van Rompay, S. Isern, S. F. Michael, L. L. Coffey, N. J. Loman, K. G. Andersen, An amplicon-based sequencing framework for accurately measuring intrahost virus diversity using PrimalSeq and iVar. Genome Biol. 20, 8 (2019).

42. P. Danecek, J. K. Bonfield, J. Liddle, J. Marshall, V. Ohan, M. O. Pollard, A. Whitwham, T. Keane, S. A. McCarthy, R. M. Davies, H. Li, Twelve years of SAMtools and BCFtools. Gigascience 10 (2021).

43. H. Wickham, M. Averick, J. Bryan, W. Chang, L. McGowan, R. François, G. Grolemund, A. Hayes, L. Henry, J. Hester, M. Kuhn, T. Pedersen, E. Miller, S. Bache, K. Müller, J. Ooms, D. Robinson, D. Seidel, V. Spinu, K. Takahashi, D. Vaughan, C. Wilke, K. Woo, H. Yutani, Welcome to the tidyverse. J. Open Source Softw. 4, 1686 (2019).

44. F. Mölder, K. P. Jablonski, B. Letcher, M. B. Hall, C. H. Tomkins-Tinch, V. Sochat, J. Forster, S. Lee, S. O. Twardziok, A. Kanitz, A. Wilm, M. Holtgrewe, S. Rahmann, S. Nahnsen, J. Köster, Sustainable data analysis with Snakemake. F1000Res. 10, 33 (2021).

45. G. M. Kurtzer, V. Sochat, M. W. Bauer, Singularity: Scientific containers for mobility of compute. PLoS One 12, e0177459 (2017).

46. K. Katoh, D. M. Standley, MAFFT Multiple Sequence Alignment Software Version 7: Improvements in Performance and Usability. Mol. Biol. Evol. 30, 772–780 (2013).

47. A. Larsson, AliView: a fast and lightweight alignment viewer and editor for large datasets. Bioinformatics 30, 3276–3278 (2014).

48. B. Q. Minh, H. A. Schmidt, O. Chernomor, D. Schrempf, M. D. Woodhams, A. von Haeseler, R. Lanfear, IQ-TREE 2: New Models and Efficient Methods for Phylogenetic Inference in the Genomic Era. Mol. Biol. Evol. 37, 1530–1534 (2020).

49. A. Rambaut, T. T. Lam, L. Max Carvalho, O. G. Pybus, Exploring the temporal structure of heterochronous sequences using TempEst (formerly Path-O-Gen). Virus Evol. 2 (2016).

50. V. Hill, G. Baele, Bayesian Estimation of Past Population Dynamics in BEAST 1.10 Using the Skygrid Coalescent Model. Mol. Biol. Evol. 36, 2620–2628 (2019).

51. A. S. Lauring, C. Edson, D. Surie, F. S. Dawood, W. H. Self, C. Lucero-Obusan, M. Holodniy, IVY Network, Genomic Characterization of RSV in the US by Vaccination Status. JAMA 333, 1540–1543 (2025).

52. Pophive, Yale School of Public Health, Population Health Information and Visualization Exchange (2025). https://github.com/PopHIVE/Ingest/raw/refs/heads/main/data/bundle_respiratory/dist/rsv_testing_pct.parquet.

53. RSV-NET, RSV hospitalization surveillance network, (2025); https://www.cdc.gov/rsv/php/surveillance/rsv-net.html.

54. D. L. Ayres, M. P. Cummings, G. Baele, A. E. Darling, P. O. Lewis, D. L. Swofford, J. P. Huelsenbeck, P. Lemey, A. Rambaut, M. A. Suchard, BEAGLE 3: Improved Performance, Scaling, and Usability for a High-Performance Computing Library for Statistical Phylogenetics. Syst. Biol. 68, 1052–1061 (2019).

55. A. J. Drummond, S. Y. W. Ho, M. J. Phillips, A. Rambaut, Relaxed Phylogenetics and Dating with Confidence. PLoS Biol. 4, e88 (2006).

56. M. A. R. Ferreira, M. A. Suchard, Bayesian analysis of elapsed times in continuous-time Markov chains. Can. J. Stat. 36, 355–368 (2008).

57. A. Rambaut, A. J. Drummond, D. Xie, G. Baele, M. A. Suchard, Posterior Summarization in Bayesian Phylogenetics Using Tracer 1.7. Syst. Biol. 67, 901–904 (2018).

58. P. Lemey, S. L. Hong, V. Hill, G. Baele, C. Poletto, V. Colizza, Á. O’Toole, J. T. McCrone, K. G. Andersen, M. Worobey, M. I. Nelson, A. Rambaut, M. A. Suchard, Accommodating individual travel history and unsampled diversity in Bayesian phylogeographic inference of SARS-CoV-2. Nat. Commun. 11, 5110 (2020).

