## Supplemental Figures 1-8; Supplemental Table 1 -4 for "Genomic surveillance of a deeply sampled local population reveals age-specific drivers of RSV transmission"

### Supplementary Materials

### Figures

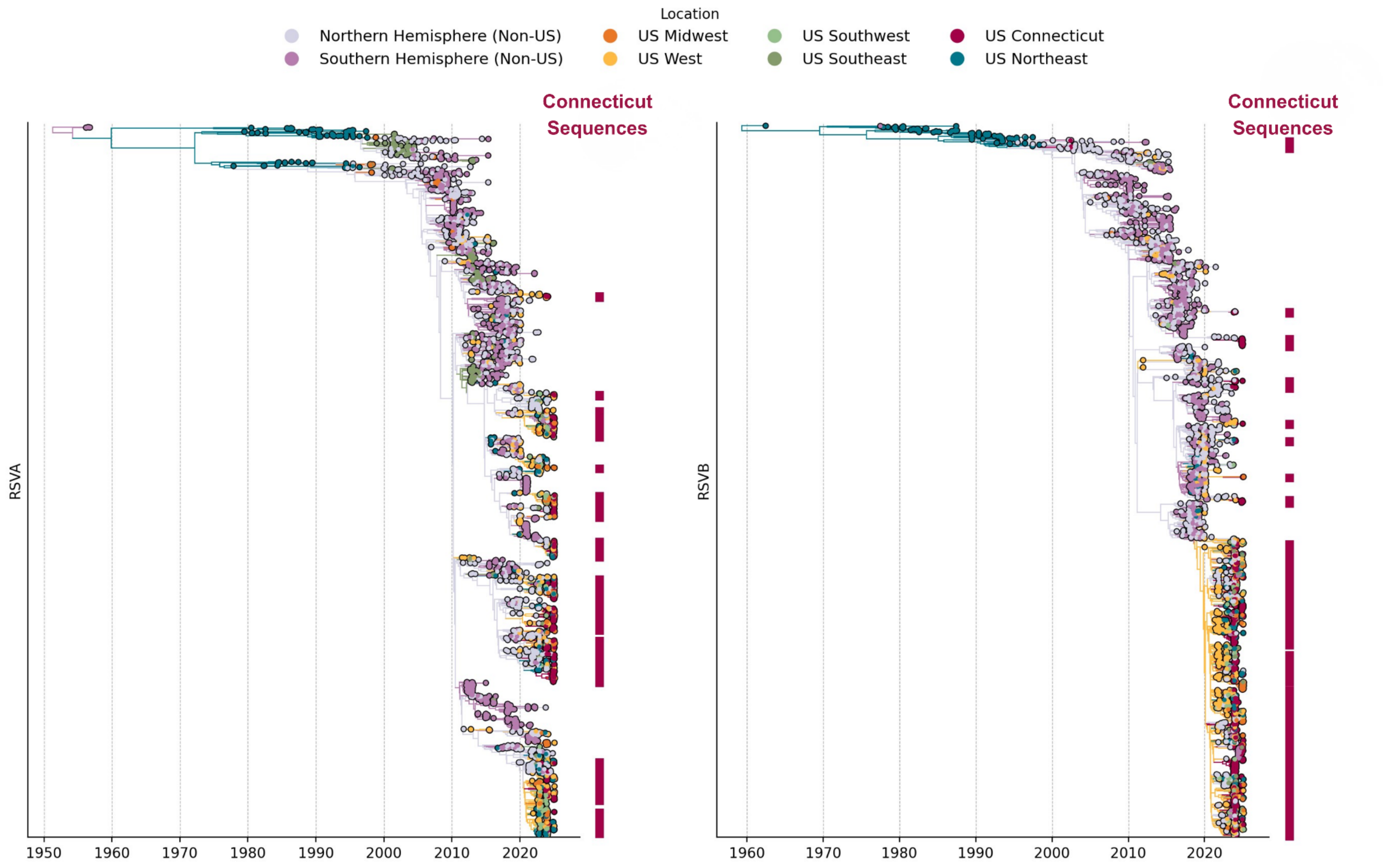


**Supplemental Figure 1. Time-resolved phylogeny of RSV-A and RSV-B tips colored by location of collection.** Lineages containing Connecticut sequences are annotated in red on the right.

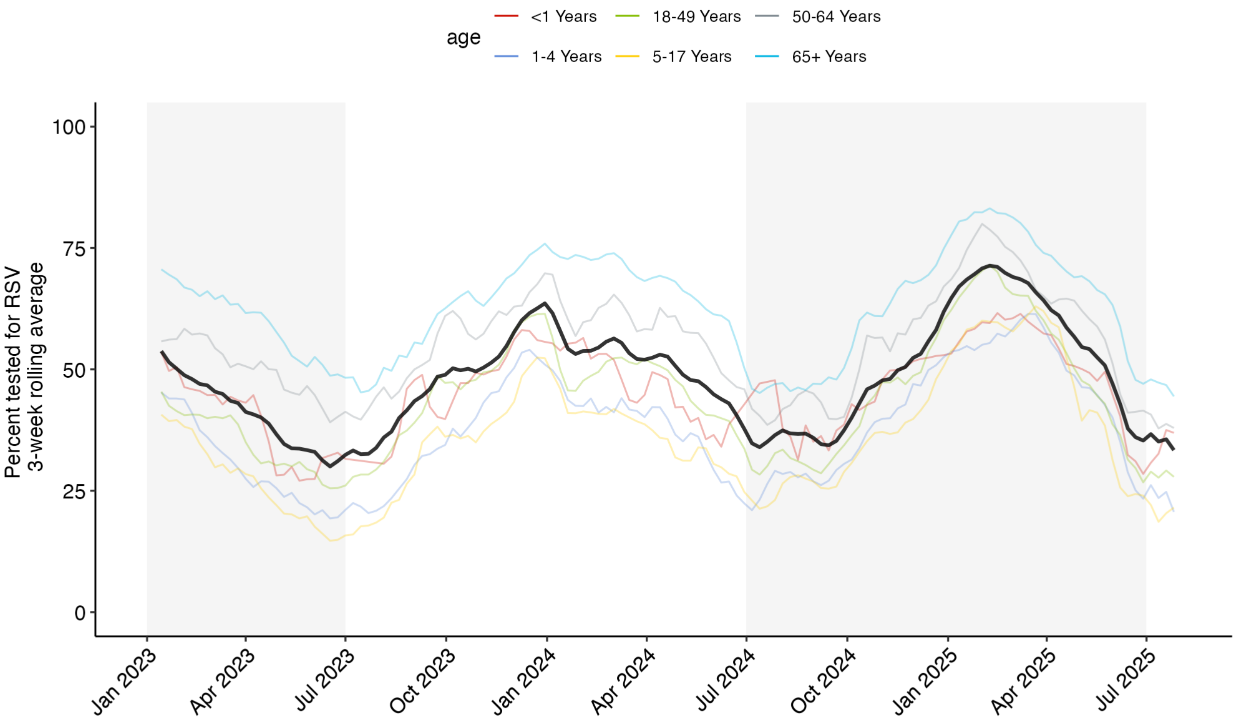


**Supplemental Figure 2. Age-stratified RSV testing rates in Connecticut, 2023-2025.** Figure displays the proportion of individuals tested for RSV, stratified by age group, with the total number of pneumonia cases as the denominator. Values are displayed as a 3-week rolling average. The solid black line represents the all-age-group 3-week rolling average RSV testing rate. These data are the result of research performed with Epic Cosmos and were obtained from the PopHIVE platform: https://github.com/PopHIVE/Ingest/blob/main/data/bundle_respiratory/dist/rsv_testing_pct.parquet.

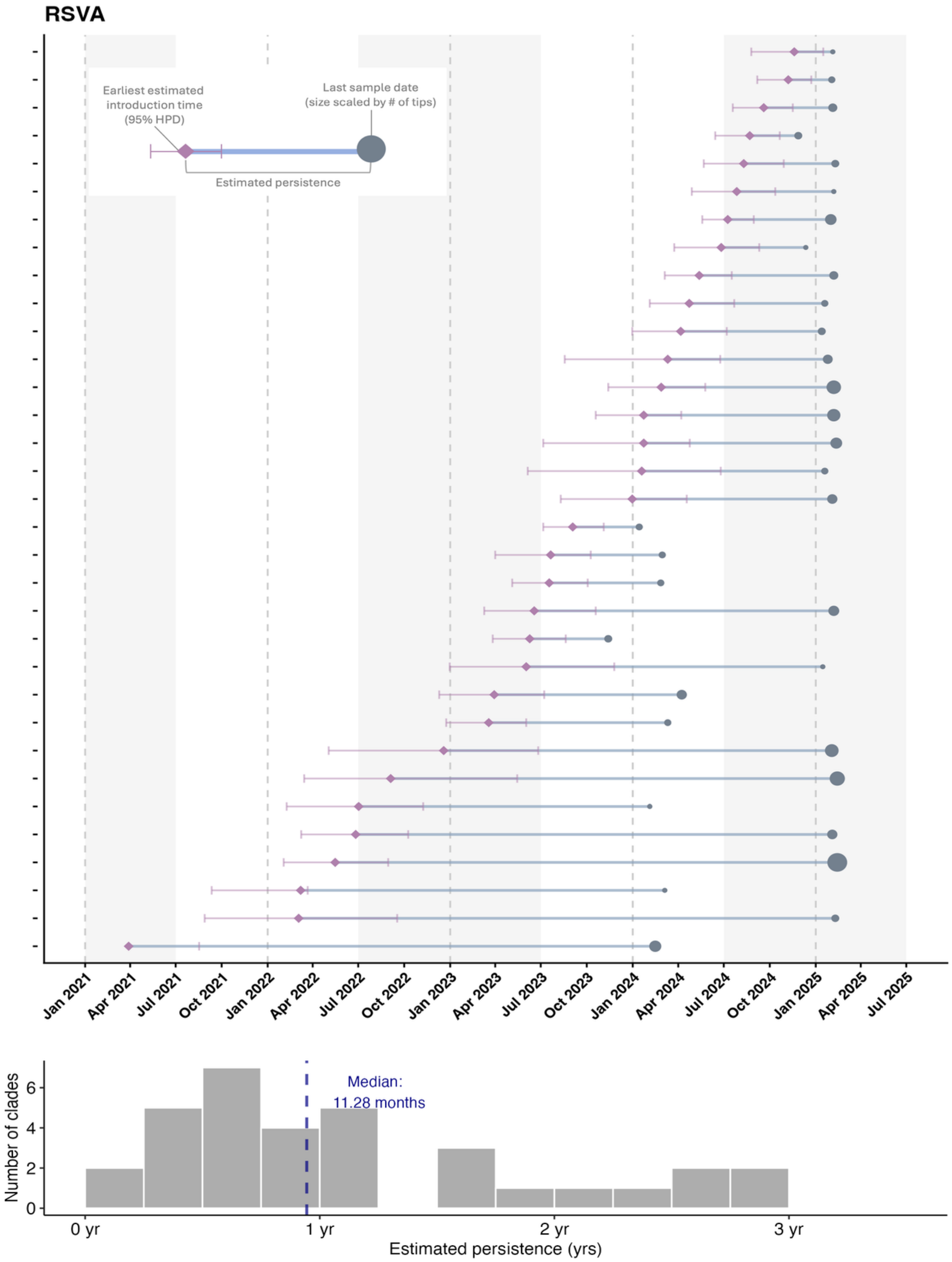


**Supplemental Figure 3. Introduction and estimated persistence of Connecticut RSV-A clades**. (A) Summary of 33 local RSV-A Connecticut clades. Each dotted line represents the start of the calendar year. (B) Histogram of estimated persistence duration. The dashed line represents the median duration.

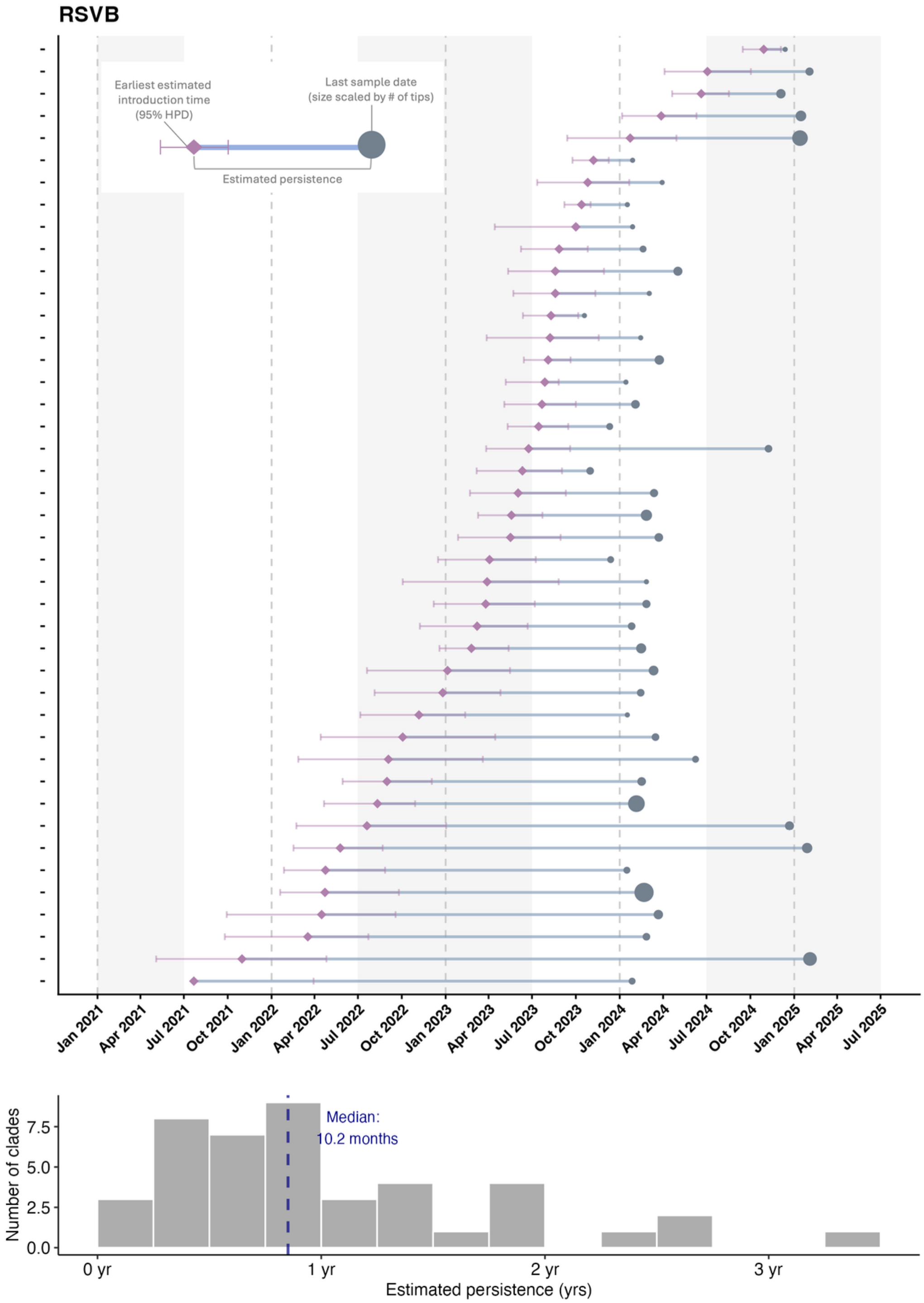


**Supplemental Figure 4. Introduction and estimated persistence of Connecticut RSV-B clades.** (A) Summary of 43 local RSV-B Connecticut clades. Each dotted line represents the start of the calendar year mark. (B) Histogram of estimated persistence duration. The dashed line represents the median duration.

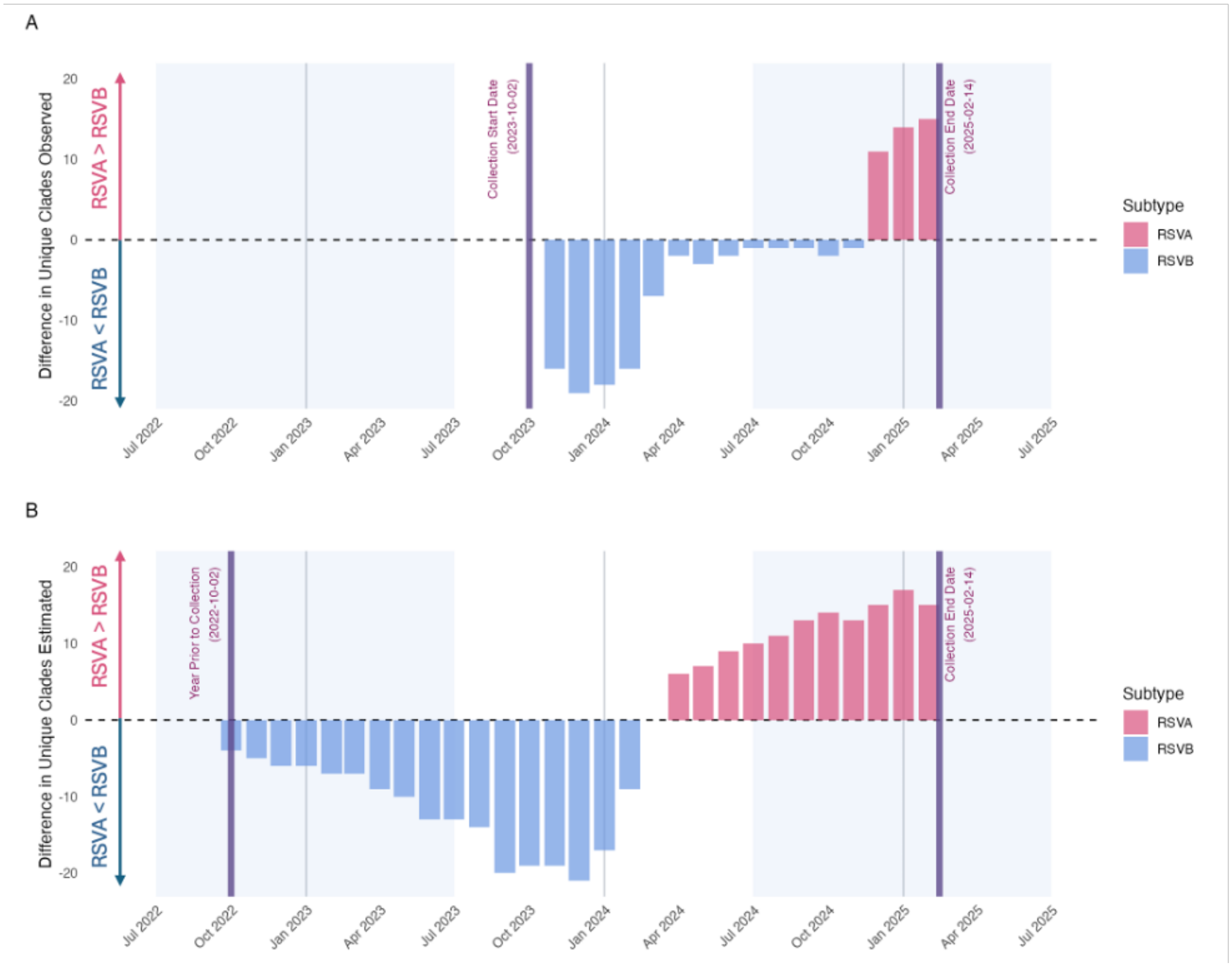


**Supplemental Figure 5. Difference in the number of unique circulating clades between RSV-A and RSV-B subtypes across two study seasons in Connecticut.** Number of circulating RSV-A clades minus RSV-B clades. (A) Observed clades, and (B) estimated clades.

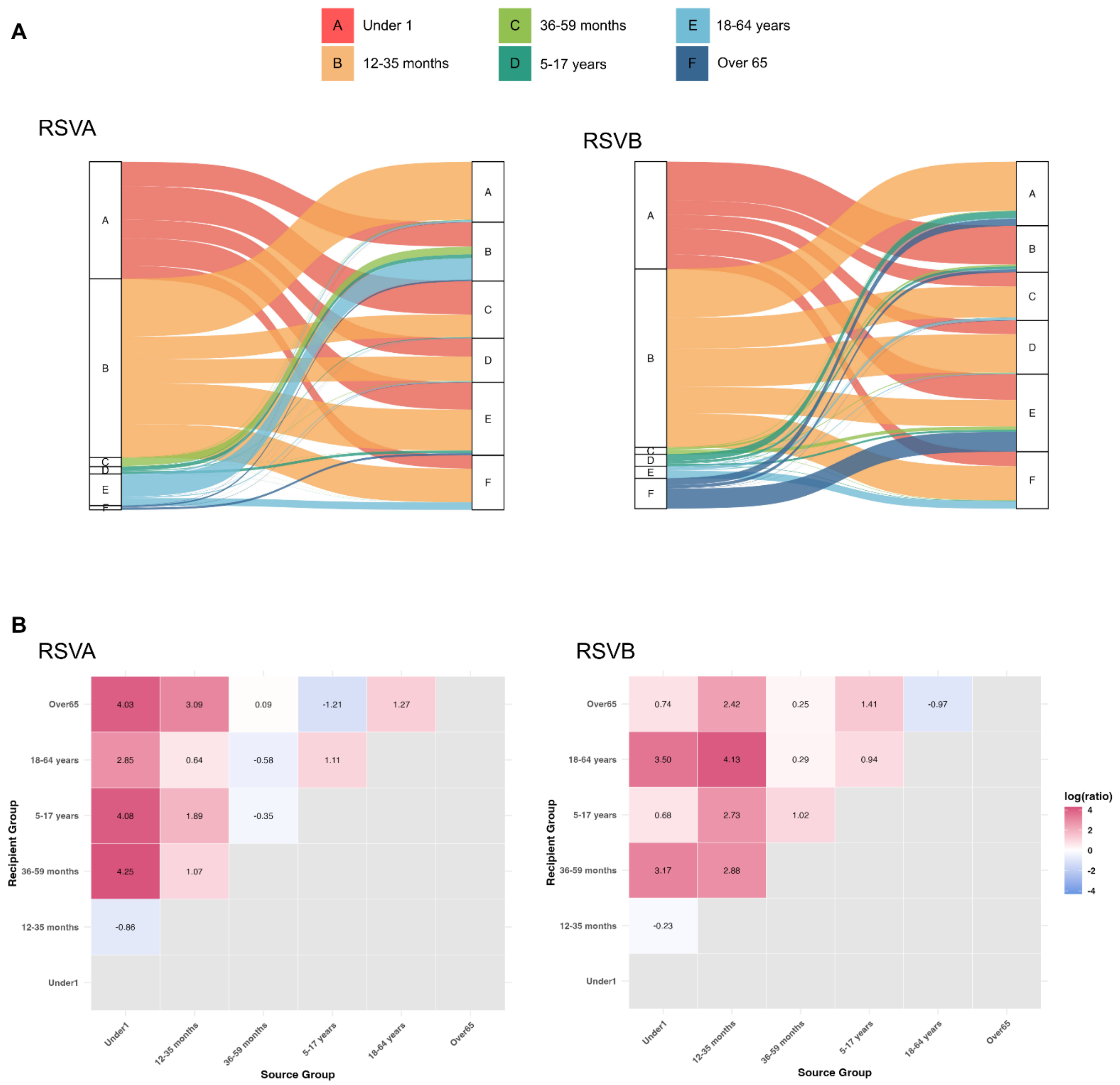


**Supplemental Figure 6. Age-stratified virus transmission summary.** (A) Sankey diagram illustrating the source and recipient flow of inter-age-group virus transmission. (B) Heatmap displaying the pairwise comparison of age groups based on source-to-recipient log ratio. White represents equal movement between the source and the recipient group. Pink shading indicates the source group is a net transmission source (ratio >1). Blue shading indicates the recipient group is a net recipient (ratio <1).

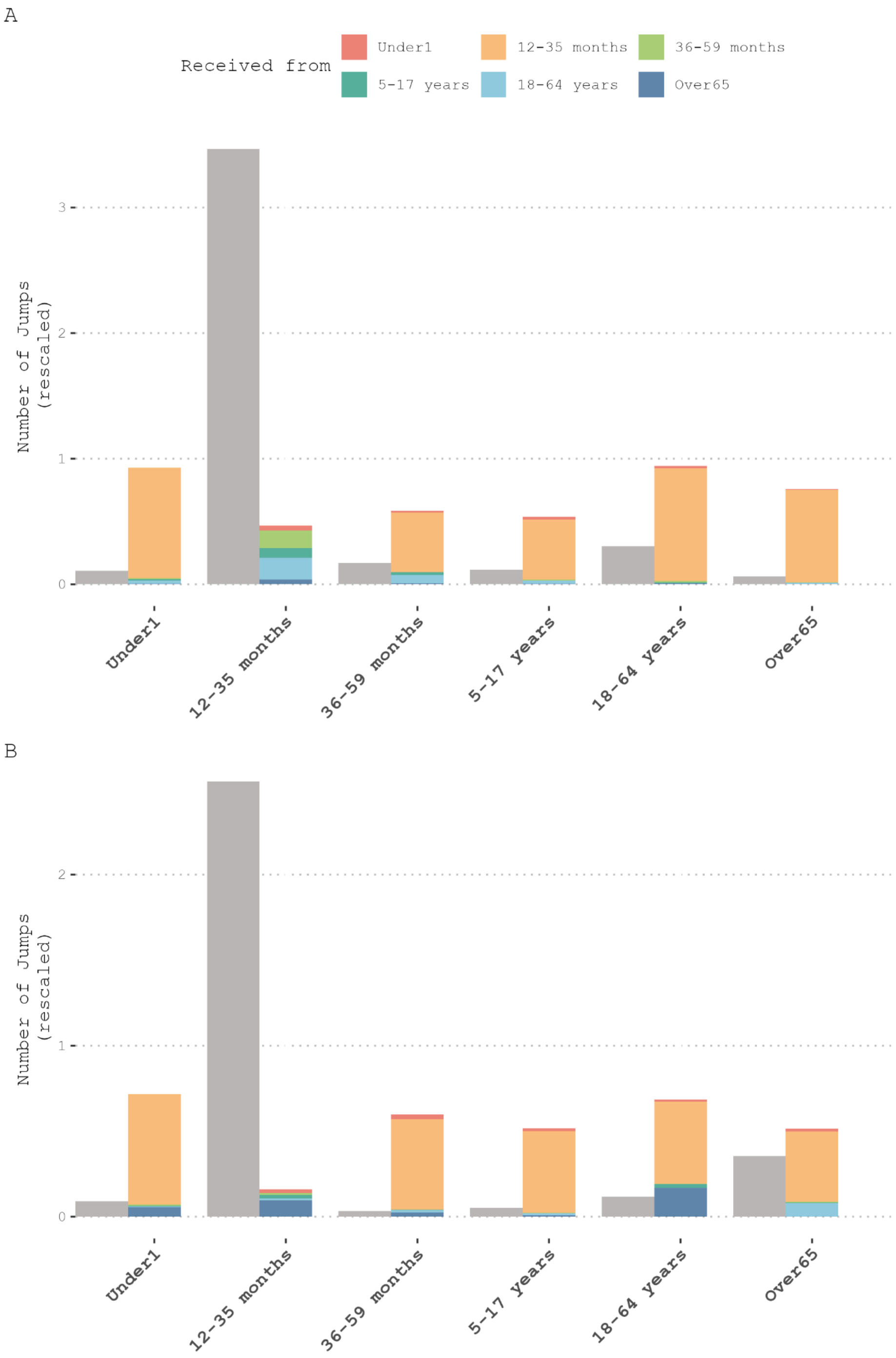


**Supplemental Figure 7**. **Sensitivity analysis (tip swap analysis) results, RSV-A (top) and RSV-B (bottom).** Age-stratified bar chart displaying the absolute number of viral transmissions given and received by each age group. Received transmissions are colored by the source age group.

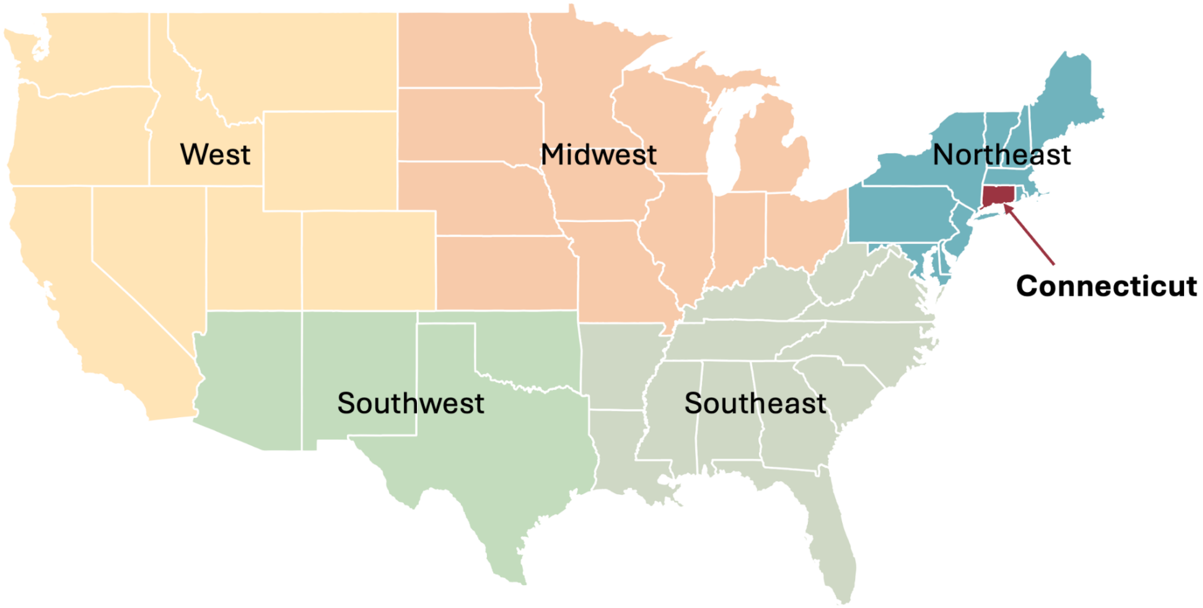


**Supplemental Figure 8.** Non-Connecticut U.S. states are mapped to one five regions: Northeast, Midwest, West, Southeast, and Southwest. Connecticut in red.

**Tables**

**Table S1. Sample distribution by age group and subtype.** Only sequences included in the discrete trait analysis.

| Age group | RSV-A, n (%) | RSV-B, n (%) |
|---|---|---|
| **Under 1** | 67 (21) | 78 (23) |
| **12-35 months** | 83 (26) | 76 (22) |
| **36-59 months** | 41 (13) | 33 (10) |
| **5-17 years** | 33 (10) | 38 (11) |
| **18-64 years** | 59 (18) | 59 (17) |
| **Over 65** | 39 (12) | 57 (17) |
| **Total** | 322 | 341 |

*Column %.

**Table S2. Sample distribution by geographic region and RSV season (Northern Hemisphere, RSVA)**

| RSV Season | Geographic Region | | | | | | | |
|---|---|---|---|---|---|---|---|---|
| | Connecticut | Midwest | Northeast | Southeast | Southwest | West | non-US NorthernH | US non-CT |
| 1997/1998 | 0 | 7 | 0 | 2 | 0 | 0 | 0 | 0 |
| 1998/1999 | 0 | 2 | 0 | 1 | 0 | 0 | 0 | 0 |
| 1999/2000 | 0 | 0 | 0 | 1 | 0 | 0 | 6 | 0 |
| 2000/2001 | 0 | 0 | 0 | 2 | 0 | 0 | 9 | 0 |
| 2001/2002 | 0 | 0 | 0 | 0 | 0 | 0 | 8 | 0 |
| 2002/2003 | 0 | 0 | 0 | 0 | 0 | 0 | 4 | 0 |
| 2003/2004 | 0 | 0 | 0 | 0 | 0 | 0 | 7 | 0 |
| 2004/2005 | 0 | 0 | 0 | 0 | 0 | 0 | 8 | 0 |
| 2005/2006 | 0 | 0 | 0 | 0 | 0 | 0 | 9 | 1 |
| 2006/2007 | 0 | 0 | 0 | 0 | 0 | 0 | 6 | 0 |
| 2007/2008 | 0 | 4 | 0 | 0 | 0 | 0 | 12 | 0 |
| 2008/2009 | 0 | 0 | 0 | 0 | 0 | 0 | 15 | 0 |
| 2009/2010 | 0 | 6 | 0 | 0 | 0 | 0 | 28 | 0 |
| 2010/2011 | 0 | 0 | 0 | 0 | 0 | 0 | 50 | 14 |
| 2011/2012 | 0 | 0 | 0 | 0 | 0 | 0 | 41 | 10 |
| 2012/2013 | 0 | 0 | 0 | 67 | 0 | 0 | 29 | 70 |
| 2013/2014 | 0 | 0 | 0 | 6 | 0 | 0 | 7 | 37 |
| 2014/2015 | 0 | 0 | 0 | 0 | 0 | 0 | 12 | 3 |
| 2015/2016 | 0 | 0 | 0 | 19 | 0 | 0 | 16 | 0 |
| 2016/2017 | 0 | 0 | 0 | 0 | 0 | 0 | 23 | 0 |
| 2017/2018 | 0 | 0 | 1 | 0 | 0 | 0 | 100 | 2 |
| 2018/2019 | 0 | 0 | 7 | 0 | 0 | 19 | 100 | 34 |
| 2019/2020 | 0 | 0 | 4 | 0 | 0 | 70 | 100 | 9 |
| 2020/2021 | 0 | 0 | 0 | 0 | 1 | 1 | 25 | 0 |
| 2021/2022 | 0 | 1 | 1 | 5 | 0 | 7 | 100 | 6 |
| 2022/2023 | 2 | 30 | 70 | 23 | 70 | 70 | 100 | 70 |
| 2023/2024 | 87 | 70 | 64 | 8 | 52 | 70 | 100 | 19 |
| 2024/2025 | 319 | 70 | 68 | 15 | 3 | 22 | 11 | 70 |

**Table S3. Sample distribution by geographic region and RSV season (Northern Hemisphere, RSVB)**

| | Geographic region | | | | | | | |
|---|---|---|---|---|---|---|---|---|
| RSV Season | Connecticut* | Midwest | Northeast | Southeast | Southwest | West | non-US NorthernH | US non-CT |
| 1998/1999 | 0 | 0 | 0 | 0 | 0 | 0 | 1 | 0 |
| 1999/2000 | 0 | 0 | 0 | 0 | 0 | 0 | 1 | 0 |
| 2000/2001 | 0 | 0 | 0 | 0 | 0 | 0 | 2 | 0 |
| 2001/2002 | 0 | 0 | 0 | 0 | 0 | 0 | 4 | 0 |
| 2002/2003 | 0 | 0 | 0 | 0 | 0 | 0 | 6 | 0 |
| 2003/2004 | 0 | 0 | 0 | 0 | 0 | 0 | 7 | 0 |
| 2004/2005 | 0 | 0 | 0 | 0 | 0 | 0 | 8 | 0 |
| 2005/2006 | 0 | 0 | 0 | 0 | 0 | 0 | 10 | 3 |
| 2006/2007 | 0 | 0 | 0 | 0 | 0 | 0 | 8 | 0 |
| 2007/2008 | 0 | 0 | 0 | 0 | 0 | 0 | 7 | 0 |
| 2008/2009 | 0 | 0 | 0 | 0 | 0 | 0 | 24 | 0 |
| 2009/2010 | 0 | 0 | 0 | 0 | 0 | 0 | 24 | 0 |
| 2010/2011 | 0 | 0 | 0 | 0 | 0 | 0 | 17 | 11 |
| 2011/2012 | 0 | 0 | 0 | 0 | 0 | 0 | 11 | 1 |
| 2012/2013 | 0 | 0 | 0 | 6 | 0 | 0 | 16 | 12 |
| 2013/2014 | 0 | 0 | 0 | 2 | 0 | 4 | 13 | 21 |
| 2014/2015 | 0 | 0 | 0 | 0 | 1 | 0 | 14 | 31 |
| 2015/2016 | 0 | 0 | 0 | 6 | 0 | 0 | 47 | 0 |
| 2016/2017 | 0 | 0 | 0 | 0 | 0 | 0 | 44 | 0 |
| 2017/2018 | 0 | 0 | 5 | 0 | 0 | 0 | 100 | 26 |
| 2018/2019 | 0 | 0 | 22 | 0 | 1 | 23 | 100 | 19 |
| 2019/2020 | 0 | 0 | 0 | 0 | 0 | 70 | 100 | 9 |
| 2020/2021 | 0 | 0 | 0 | 0 | 0 | 4 | 45 | 1 |
| 2021/2022 | 0 | 0 | 0 | 0 | 1 | 70 | 100 | 55 |
| 2022/2023 | 0 | 3 | 19 | 6 | 28 | 70 | 100 | 30 |
| 2023/2024 | 385 | 70 | 70 | 31 | 68 | 70 | 100 | 70 |
| 2024/2025 | 98 | 70 | 31 | 17 | 6 | 19 | 3 | 51 |

* Three sequences from 2002 with month of collection unknown.

**Table S4. Sample distribution by subtype and RSV season (Southern Hemisphere).** Including sequences with unknown months of collection.

| Year | RSV Subtypes | |
|---|---|---|
| | RSVA | RSVB |
| 1956 | 3 | 0 |
| 1977 | 0 | 1 |
| 2002 | 0 | 1 |
| 2004 | 3 | 2 |
| 2005 | 1 | 0 |
| 2006 | 3 | 2 |
| 2007 | 10 | 6 |
| 2008 | 37 | 9 |
| 2009 | 49 | 28 |
| 2010 | 110 | 104 |
| 2011 | 19 | 16 |
| 2012 | 101 | 11 |
| 2013 | 38 | 13 |
| 2014 | 71 | 59 |
| 2015 | 77 | 59 |
| 2016 | 85 | 104 |
| 2017 | 100 | 110 |
| 2018 | 129 | 128 |
| 2019 | 100 | 100 |
| 2020 | 100 | 20 |
| 2021 | 88 | 9 |
| 2022 | 69 | 14 |
| 2023 | 21 | 24 |
| 2024 | 1 | 56 |
